# A simple criterion to design optimal non-pharmaceutical interventions for epidemic outbreaks

**DOI:** 10.1101/2020.05.19.20107268

**Authors:** Marco Tulio Angulo, Fernando Castaños, Rodrigo Moreno-Morton, Jorge X. Velasco-Hernández, Jaime A. Moreno

## Abstract

For mitigating the COVID-19 pandemic, much emphasis is made on implementing non-pharmaceutical interventions to keep the reproduction number below one. However, using that objective ignores that some of these interventions, like bans of public events or lockdowns, must be transitory and as short as possible because of their significative economic and societal costs. Here we derive a simple and mathematically rigorous criterion for designing optimal transitory non-pharmaceutical interventions for mitigating epidemic outbreaks. We find that reducing the reproduction number below one is sufficient but not necessary. Instead, our criterion prescribes the required reduction in the reproduction number according to the desired maximum of disease prevalence and the maximum decrease of disease transmission that the interventions can achieve. We study the implications of our theoretical results for designing non-pharmaceutical interventions in 16 cities and regions during the COVID-19 pandemic. In particular, we estimate the minimal reduction of each region’s contact rate necessary to control the epidemic optimally. Our results contribute to establishing a rigorous methodology to guide the design of optimal non-pharmaceutical intervention policies.

## Introduction

Since the seminal work of May and Anderson [1], the design of interventions to *eradicate* infectious diseases has the objective of achieving a basic (*R*_0_) or effective reproduction number below one [2, 3]. The underlying assumption here is that it is possible to maintain interventions for long periods, such as long-term vaccination programs. During the COVID-19 pandemic, this same objective is guiding the design of non-pharmaceutical interventions (NPIs) [4]. However, maintaining NPIs like bans of public events or lockdowns for long periods of time is infeasible because of their substantial economic and societal costs [5, 6]. Actually, instead of aiming for eradication, NPIs aim to *mitigate* the economic and social costs of the epidemic outbreak [7]. Nevertheless, we still lack simple guidelines to design NPIs for mitigating epidemic outbreaks, analogous to the *R*_0_ < *1* condition for eradication.

Here we use the classical Susceptible-Infected-Removed epidemiological model to fully characterize the design of NPIs for mitigating epidemic outbreaks. With this aim, we consider that NPIs should achieve an optimal tradeo? between two objectives [8]. First, optimal NPIs must minimize the period in which they need to be applied, consequently minimizing their associated economic and societal costs. Second, optimal NPIs must guarantee that the disease prevalence does not exceed a specified maximum level, which for example can represent the health services’ capacity for that particular disease [9]. We obtain a full analytical characterization of such optimal NPIs, specifying the optimal intervention at each state that the epidemic can be. This characterization yields the necessary and sufficient criterion for the existence of optimal NPIs for mitigation, analogous to the *R*_0_ < *1* condition for eradication. We find that reducing the reproduction number below one is sufficient but not necessary for their existence. Instead, we show that the desired maximum disease prevalence determines the necessary reduction in the reproduction number. The consequence of not reducing the reproduction number below one is that interventions must start before the disease prevalence reaches the specified maximum level. We also demonstrate numerically that the derived optimal NPIs are robust to uncertainties in the model parameters and unmodeled epidemic dynamics (e.g., undetected infections). Finally, we explore the implications of our theoretical result by analyzing the response of 16 cities and regions across the globe to the COVID-19 pandemic, finding that most regions achieved a larger-than-necessary reduction in transmission. Our results contribute to designing non-pharmaceutical interventions to respond optimally and robustly against epidemic outbreaks.

## Characterizing optimal non-pharmaceutical interventions

### Optimal epidemic mitigation using NPIs

Our objective is to characterize the reduction in the disease transmission that is optimal for each *state* in which the epidemic outbreak can be. For this, we leverage on the mathematical tractability of the Susceptible-Infected-Removed (SIR) model [10], where the state can be characterized by the pair (*S, I*) ∈ [0, 1]^2^. Here, *S* is the proportion of the population that is susceptible to the disease, and *I* is the disease prevalence (i.e., proportion of the population that is infected), see Fig. 1a. We discuss later other more detailed epidemic models. The epidemic state changes with time *t* as the disease is transmitted, producing the trajectory (*S*(*t*), *I*(*t*)) for *t* ≥ 0. For epidemic *mitigation*, we consider that the goal is keeping the disease prevalence below a specified level *I*_max_ 2 (0, 1]. This constant may characterize, for example, the health services’ capacity in the sense that a prevalence above *I*_max_ causes higher mortality due to hospital saturation [11]. In general, *I*_max_ should consider all social and economic conditions of the specific population where the outbreak occurs. To keep *I*(*t*) ≤ *I*_max_, we assume we can apply one or several NPIs that reduce disease transmission by the factor (1 − *u*), for some *u* ∈ [0, 1], see Fig. 1a. The NPIs achieve no reduction when *u* = 0, and they completely stop transmission when *u* = 1. Since it is unfeasible to stop transmission fully, we upper-bound the reduction by *u*_max_ ∈ (0, 1). We say that *u* is *admissible* if *u* ∈ [0, *u*_max_].

**Figure 1:**
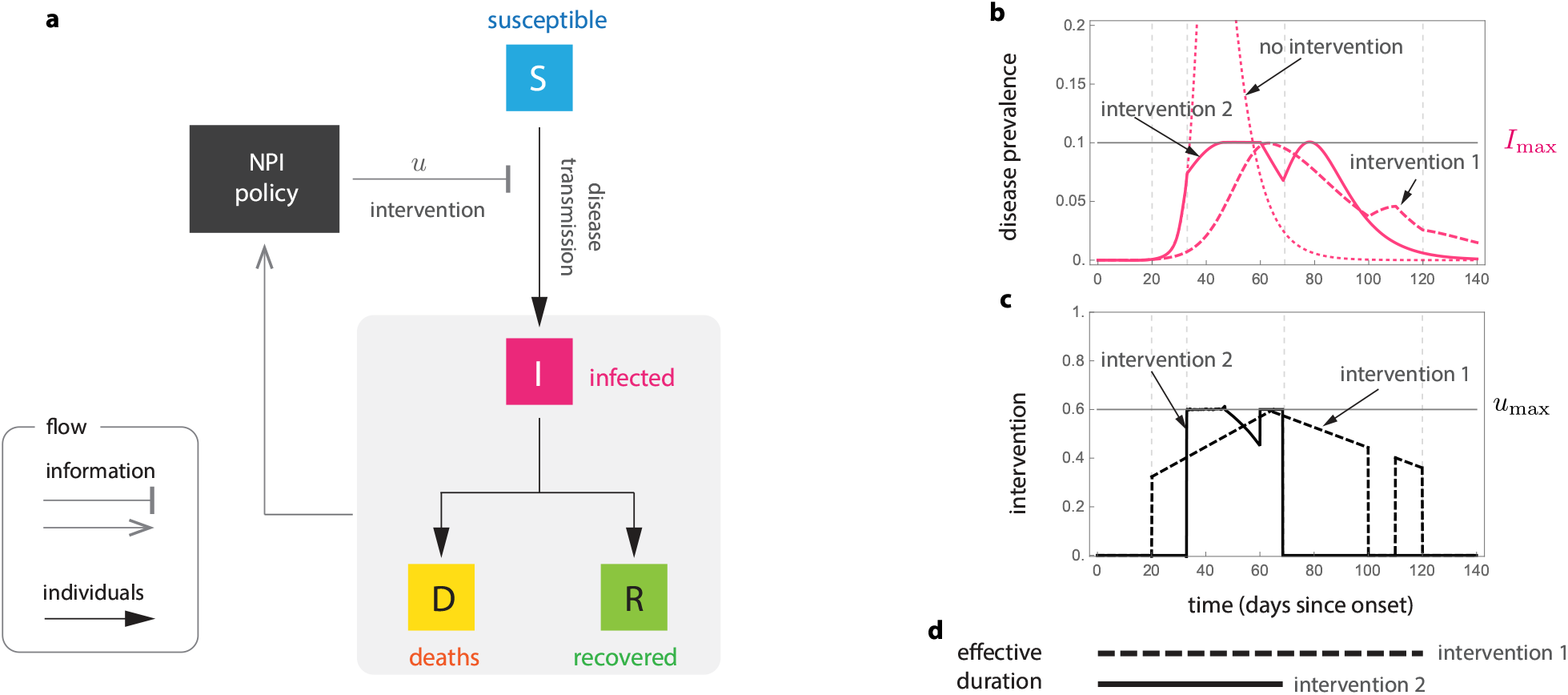
Optimal non-pharmaceutical interventions. **a**. Susceptible-Infected-Removed (SIR) model with non-pharmaceutical interventions (NPIs) reducing disease transmission. For the optimal NPI design problem, the objective is to design the intervention *u*^∗^(*t*) with minimal effective duration such that *u*^∗^(*t*) ∈ [0, *u*_max_] and *I*(t) ≤ *I*_max_ for all *t* ≥ 0. **b and c**. Panels show the response of the SIR model for two interventions (parameters are *β* = 0.52, *γ* = 1/7, *I*_0_ = 8.855 x 10^−7^ and *S*_0_ = 1 − *I*_0_). Both intervention 1 and 2 satisfy *u*(*t*) ≤ *u*_max_ and guarantee that *I*(*t*) ≤ *I*_max_ Actually, intervention 2 is the optimal one derived using our analysis: it is intervention with minimal effective duration satisfying *I*(*t*) ≤ *I*_max_. **d**. The effective duration of an intervention measures the interval between the start of the outbreak and the last time that a non-zero intervention is applied. In this example, the effective duration of intervention 1 is 120 days, while the effective duration of intervention ∈ is 69 days.

Different admissible NPIs can keep the disease prevalence below *I*_max_. For instance, “intervention 1” in the example of Fig. 1b-c keeps this restriction and has an “effective duration” of 120 days. Here, the *effective duration* of an intervention is the interval between the start of the outbreak and the last time that a non-zero intervention is applied (Fig. 1d). “Intervention 2” of Fig. 1b-c also keeps the restriction *I*(*t*) ≤ *I*_max_, but its effective duration is only 69 days. To design the *optimal* NPI, we ask for the intervention with minimal effective duration. Specifically, we ask for the admissible reduction *u*^∗^ (*S*(*t*), *I*(*t*)) required *now* (i.e., at the current state) such that: (1) it minimizes the effective duration of the intervention; and (2) it ensures that the prevalence can be maintained below *I*_max_ for *all future* time by using some admissible intervention. If the optimal NPI problem has a solution *u*^∗^, then *u*^∗^(*S, I*) characterizes the optimal reduction in the disease transmission that the NPIs should achieve if the epidemic state is (*S, I*). In particular, *u*^∗^ gives the optimal way to start and stop the NPIs.

### Optimal NPIs exist without reducing the reproduction number below one

Our first main result is a complete analytical characterization of the optimal NPIs in the SIR model (see Box 1 for a summary and Supplementary Note S1 for details). To understand how the optimal NPIs work, note that the SIR model predicts a *safe zone* of states (*S, I*) where, without any further interventions, the disease prevalence will not exceed *I*_max_ (blue zone in Fig. 2a-c). The safe zone is characterized by the inequality 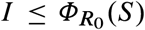, where *R*_0_ is the *basic reproduction number* of the outbreak in the population, and the function *Φ*_*R*_ is defined in Eq. (2) of Box 1. The goal of an optimal NPI is thus to reach this safe zone as fast as possible without violating the restriction *I*(*t*) ≤ *I*_max_. The ability to achieve this goal depends on the epidemic state. That is, we can partition the plane (*S, I*) in two regions: those states from which it is possible to reach the safe zone without exceeding *I*_max_ (*feasible* states), and those where it is impossible (*unfeasible* states). We find these two regions are characterized by the separating curve 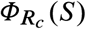, where we call *R*_*c*_ := (1 − *u*_max_)*R*_0_ the *controlled reproduction number* (Fig. 2a-c). Note that *R*_*c*_ describes the maximum reduction in the basic reproduction number that (constant) admissible interventions can achieve. Therefore, *R*_*c*_ < 1 is the necessary and sufficient condition that a constant and permanent admissible intervention (i.e., *u*(*t*) ≡ const. for all *t* ≥ 0) needs to satisfy to *eradicate* a disease outbreak in the SIR model. However, for outbreak mitigation, our analysis shows that feasible states exists without achieving disease eradication (white regions in Fig. 2b-c). This result is important because it proves that optimal NPIs for epidemic mitigation do not require reducing the basic reproduction number below one.

**Figure 2:**
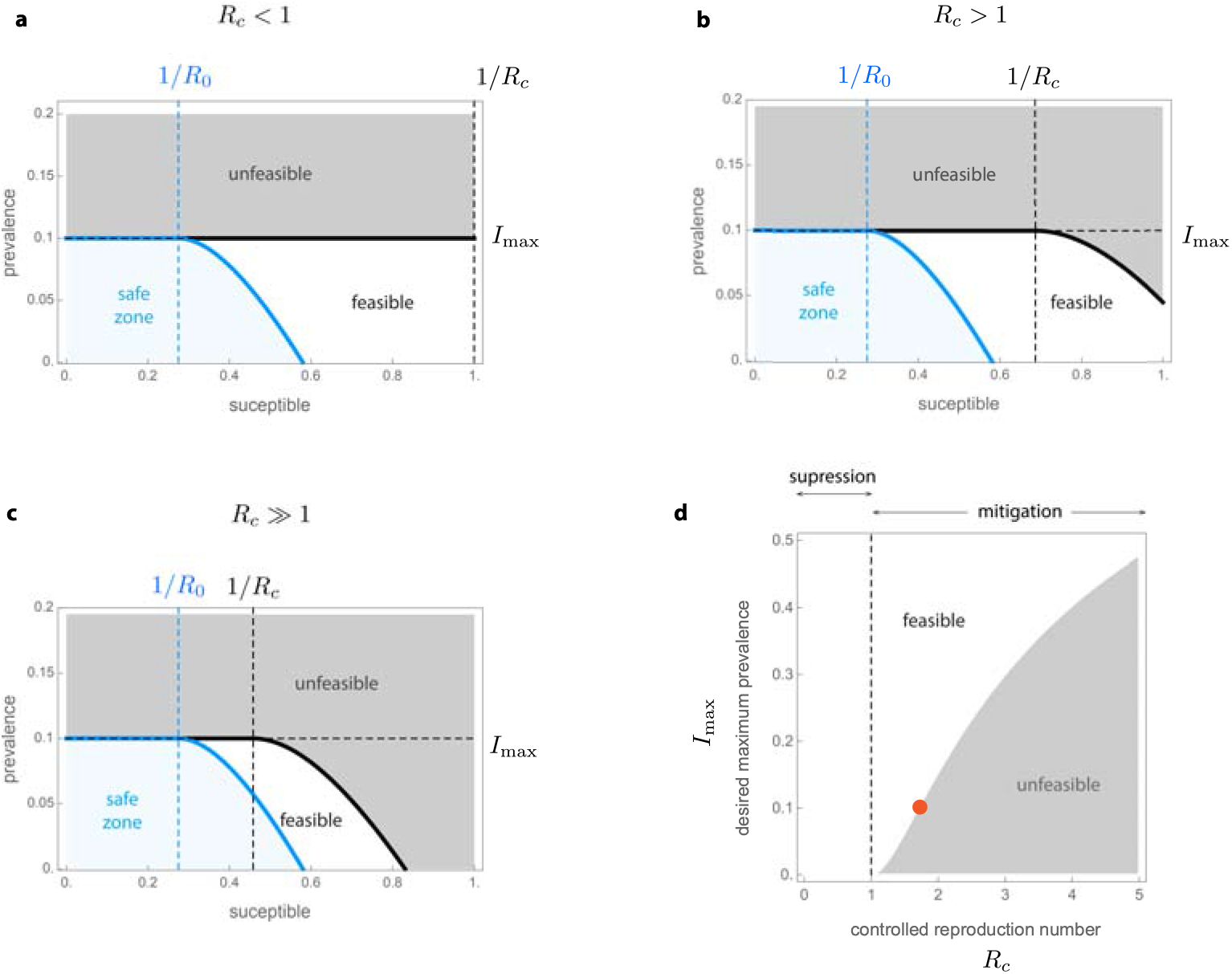
Existence of non-pharmaceutical interventions in the Susceptible-Infected-Removed model. Parameters are *γ* = 1/7, *β* = 0.52, (i.e., *R*_0_ = 3.64) and *I*_max_ = 0.1. The safe zone (in blue) consists of all states that dot not exceed *I*_max_ without interventions. This zone is characterized by the inequality 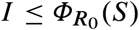. The plane is further divided into feasible states that can reach the safe zone without exceeding *I*_max_ (white), and unfeasible states that cannot (gray). Feasible and unfeasible states are separated by the separating curve 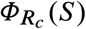 (black line). **a**. For “strong” interventions with *u*_max_ = 0.8, the controlled reproduction number is *R*_*c*_ = (1 − *u*_max_)*R*_0_ = 0.728 < 1. Here, the separating curve is the straight line *I*_max_, implying that all states below *I*_max_ are feasible. Note this case corresponds to eradication. **b**. For “intermediate” interventions with *u*_max_ = 0.6, the controlled reproduction number is *R*_*c*_ = (1−*u*_max_)*R*_0_ = 1.456 > 1. Here, the separating curve 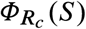 is nonlinear, and some states below *I*_max_ are unfeasible. **c**. For “weak” interventions with *u*_max_ = 0.4 we obtain *R*_*c*_ = 2.184 > 1. In this case, states with *S*(0) ≈ 1 are unfeasible. **d**. For *S*(0) → 1, our design criterion for NPIs prescribe the values of *R*_*c*_ ‘s that a given *I*_max_ can manage.

### A design criterion for optimal NPIs

We demonstrated above that optimal NPIs exist even when *R*_*c*_ > 1. However, how large can *R*_*c*_ be before NPIs keeping *I*(*t*) ≤ *I*_max_ do not exist? When *S*(0) → 1, our characterization shows that an NPIs exists if and only if

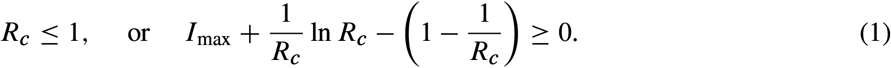

The above inequality is our second main result, connecting the specified maximum disease prevalence *I*_max_ with the outbreak’s controlled reproduction number *R*_*c*_ = (1 − *u*_max_)*R*_0_ (Supplementary Note S2). The inequality (1) governs the existence of NPIs for mitigating epidemic outbreaks, in analogy to how the condition *R*_*c*_ < 1 works for disease eradication. Note that *R*_*c*_ < 1 is a sufficient condition for the existence for NPIs, but the inequality (1) shows that this condition is far from necessary. If *I*_max_ > 0, there exists *R*_*c*_ > 1 for which NPIs exist (Fig. 2d). Note also that the maximum feasible *R*_*c*_ increases with *I*_max_.

We can use (1) to design NPIs as follows. Consider an infectious disease outbreak with a given *R*_0_ and that the specified maximum prevalence is *I*_max_. Then, the inequality (1) gives the criterion to design NPIs by providing the range of disease transmission reduction *u*_max_ that the NPIs should attain. In particular, it provides the minimal reduction 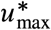 in the contact rate required for the existence of NPIs. For example, if *I*_max_ = 0.1 then 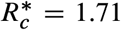 is the maximum admissible controlled reproduction number (orange point in Fig. 2d). Therefore, if an outbreak in the population has *R*_0_ = 3, then the minimal reduction is 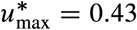 because 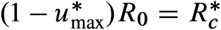.

### Optimal NPIs are simple

For any epidemic state, the optimal transmission reduction takes a simple form which can be described by coloring the (*S, I*) plane, see top row of Fig. 3. Here, for all states in the white region the optimal intervention is no intervention; for all states in the yellow region the optimal intervention is *u*^∗^(*S, I*) = *u*_max_. There are regions (specifically lines) where the optimal intervention switches frequently between *u*^∗^ = 0 and *u*^∗^ = *u*_max_ producing a so-called “singular arc” that slides along the two regions, leading to an “average” intervention *u*^∗^ ∈ [0, *u*_max_]. We find that, in general, the optimal NPIs have four phases: a first one where no intervention is needed, a second phase where interventions start with maximum strength, a third phase of gradual decrease of interventions, and a “final push” where the maximum interventions are re-applied for a short period to reach the safe zone faster.

**Figure 3:**
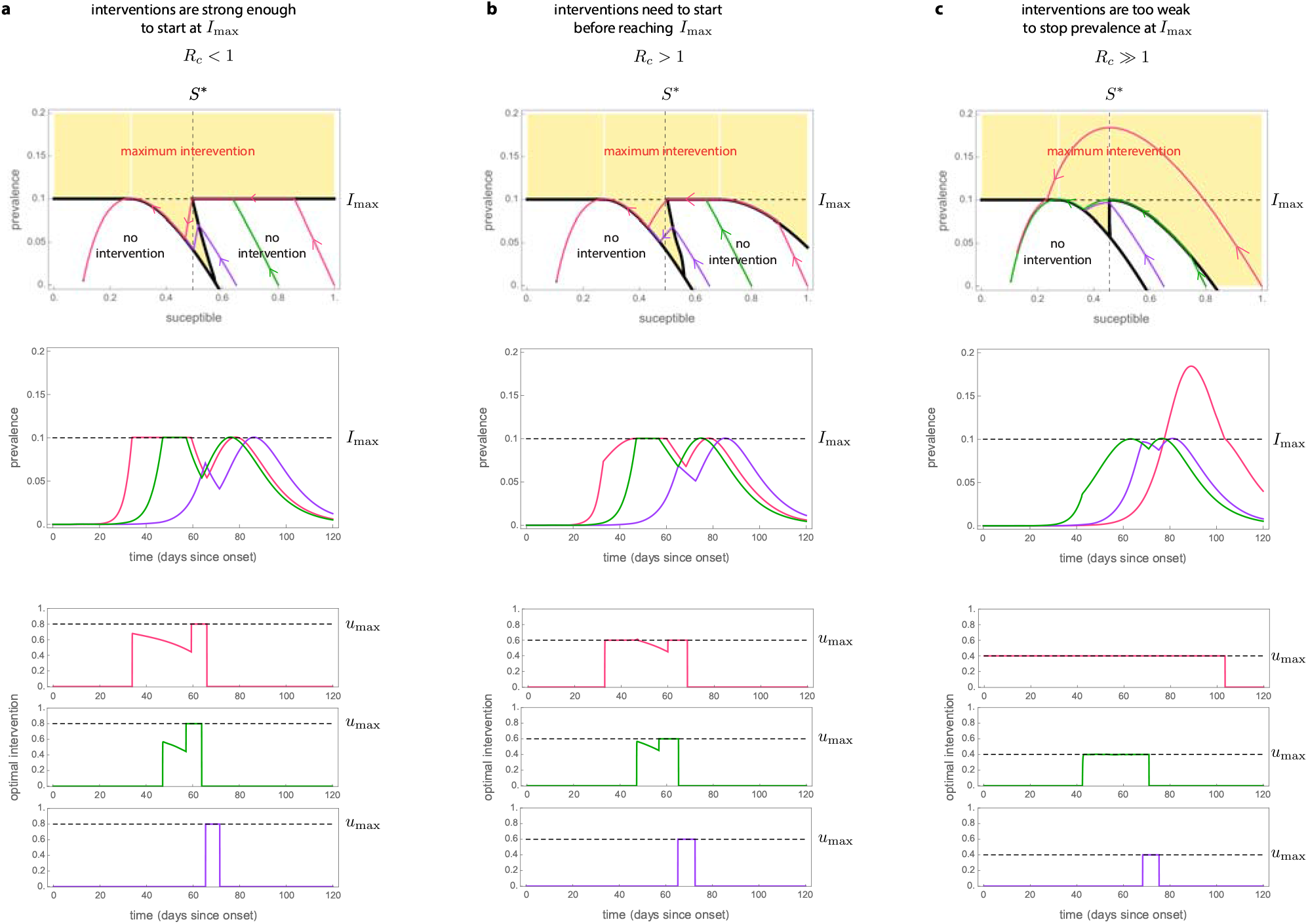
Optimal non-pharmaceutical interventions in the Susceptible-Infected-Removed model. For all panels, the parameters of the SIR model are *γ* = 1/7, *β* = 0.52, (i.e., *R*_0_ = 3.64) and *I*_max_ = 0.1. We consider a population of *N* = 8.855 x 10^6^ individual (like in Mexico City) and *I*_0_ = 1*/N*. Panels shows trajectories for three initial proportions of the susceptible population: large *S*_0_ = 1 − *I*_0_ ≈ 1 (pink), medium *S*_0_ = 0.8 (green), and small *S*_0_ = 0.65 (purple). **a**. For *u*_max_ = 0.8 we have *R*_*c*_ = (1 − *u*_max_)*R*_0_ = 0.728 ≤ 1. In this case, the optimal intervention starts when the disease prevalence reaches *I*_max_. Afterwards, the intervention decreases in an hyperbolic arc until reaching the point *S* = *S*^∗^. At that time, the intervention becomes maximum in the “final push” to reach the safe zone. **b**. For *u*_max_ = 0.58 the controlled reproduction number is *R*_*c*_ = (1 − *u*_max_)*R*_0_ = 1.52 > 1. Here 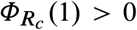, implying that the epidemic still can be mitigated for initial states with *S*_0_ ≈ 1 and *I*_0_ ≈ 0 (pink trajectory). In this case, the optimal intervention starts when the initial condition hits the separating curve below *I*_max_ at *t* = 35. At that instant the intervention starts with the maximum value *u*_max_, and continues in that form until the trajectory reaches *I*_max_. **c**. Choosing *u*_max_ = 0.4 yields *R*_*c*_ = 2.184 > 1. In this case, the optimal intervention problem does not have a solution for all initial states *S*_0_ > 0.85. This is illustrated by pink trajectory: even when applying the maximum intervention from the start, *I*(*t*) will grow beyond *I*_max_.

We illustrate the above behavior in three qualitatively Different cases. The first case is when the optimal intervention starts just when the disease prevalence reaches *I*_max_ (Fig. 3a). This case occurs when the interventions are strong enough to stop the rise in prevalence at *I*_max_ regardless of the fraction of susceptible population. Our analysis shows that this occurs if and only if *u*_max_ is large enough to render *R*_*c*_ = (1 − *u*_max_)*R*_0_ ≤ 1. When the initial susceptible population is close to 1 (pink trajectory in Fig. 3a), the optimal intervention first waits until the disease prevalence reaches *I*_max_. At that time, the optimal NPI stops the disease prevalence exactly at *I*_max_, and then it gradually decreases its magnitude to ensure that the disease prevalence slides along *I*_max_ as the susceptible population decreases. When the susceptible population reaches the threshold *S*^∗^, the optimal intervention is again the maximum one (Fig. 3a). This “final push” allows reaching the safe zone faster, releasing the interventions sooner. The middle and bottom panels of Fig. 3a show the resulting disease prevalence and optimal interventions as a function of time. Note that a smaller initial susceptible population yields other trajectories (green and purple in Fig. 3a).

The second case is when an “early” intervention is necessary before the disease prevalence reaches *I*_max_ (Fig. 3b). This case happens when the admissible reduction in the contact rate cannot immediately stop the disease prevalence at *I*_max_ if the susceptible population is large at that time. We find this case occurs if and only if *u*_max_ is small in the sense that *R*_*c*_ = (1 − *u*_max_)*R*_0_ > 1. Here, a trajectory may hit the yellow region before reaching *I*_max_ (pink trajectory in Fig. 3b). When that happens, the optimal intervention starts with the maximum reduction *u*^∗^ = *u*_max_. Then it maintains this maximum reduction to “slide” the trajectory between the yellow and white regions. Once the trajectory reaches *I*_max_, the magnitude of the optimal intervention decreases to slide the trajectory along *I*_max_. Again, the final push occurs when the susceptible population reaches *S*^∗^.

The third case is when the initial state (*S*_0_; *I*_0_) lies in the unfeasible region (Fig. 3c). This case occurs when *u*_max_ is so small that, even if the maximum admissible intervention *u* = *u*_max_ is applied from the start of the outbreak, the disease prevalence will exceed *I*_max_ (pink trajectory in Fig. 3c). In this case the optimal intervention problem is unfeasible because it is impossible to achieve *I*(*t*) ≤ *I*_max_. However, note that the using *u*^∗^ = *u*_max_ yields the smallest prevalence peak.

### Optimal NPIs are robust

To evaluate the optimal NPIs in more realistic scenarios, we numerically analyzed their performance in three epidemic models with uncertain epidemic parameters and more detailed epidemic dynamics (see details in Supplementary Note S3). In all cases, we consider that the basic reproduction number has been estimated as 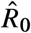 using an SIR model, and that the optimal NPIs are designed using this estimate. Then, these optimal NPIs are applied to an outbreak with possibly Different epidemic dynamics and possibly Different *R*_0_. Note that estimation errors in *R*_0_ will affect the correct start and “final push” for reaching the safe zone.

In the first scenario, we consider an outbreak with SIR dynamics where the strength of the NPIs is uncertain. We model this uncertainty replacing *u* by *ku* in the model equations, where *k* ∈ (0, 1). Then, for example, *k* = 0.9 (resp. *k* = 1.1) represents a 10% underestimation (resp. overestimation) of the NPIs strength. Across outbreaks with Different *R*_0_’s and an uncertainty of 10% in the intervention’s strength, we find that the disease prevalence is maintained below *I*_max_ as long as *R*_0_ is not underestimated (Fig. 4a). In the second scenario, we consider an SEIR outbreak with an incubation period for the disease. For an incubation period of 7 days as in a typical COVID-19 infection, the optimal NPIs maintain the disease prevalence below *I*_max_ if *R*_0_ < 2.5 and its value is estimated with an error of below 30% (solid yellow and orange in Fig. 4b). For larger *R*_0_ or a larger incubation period, the disease prevalence may exceed *I*_max_ (red in Fig. 4b).

**Figure 4:**
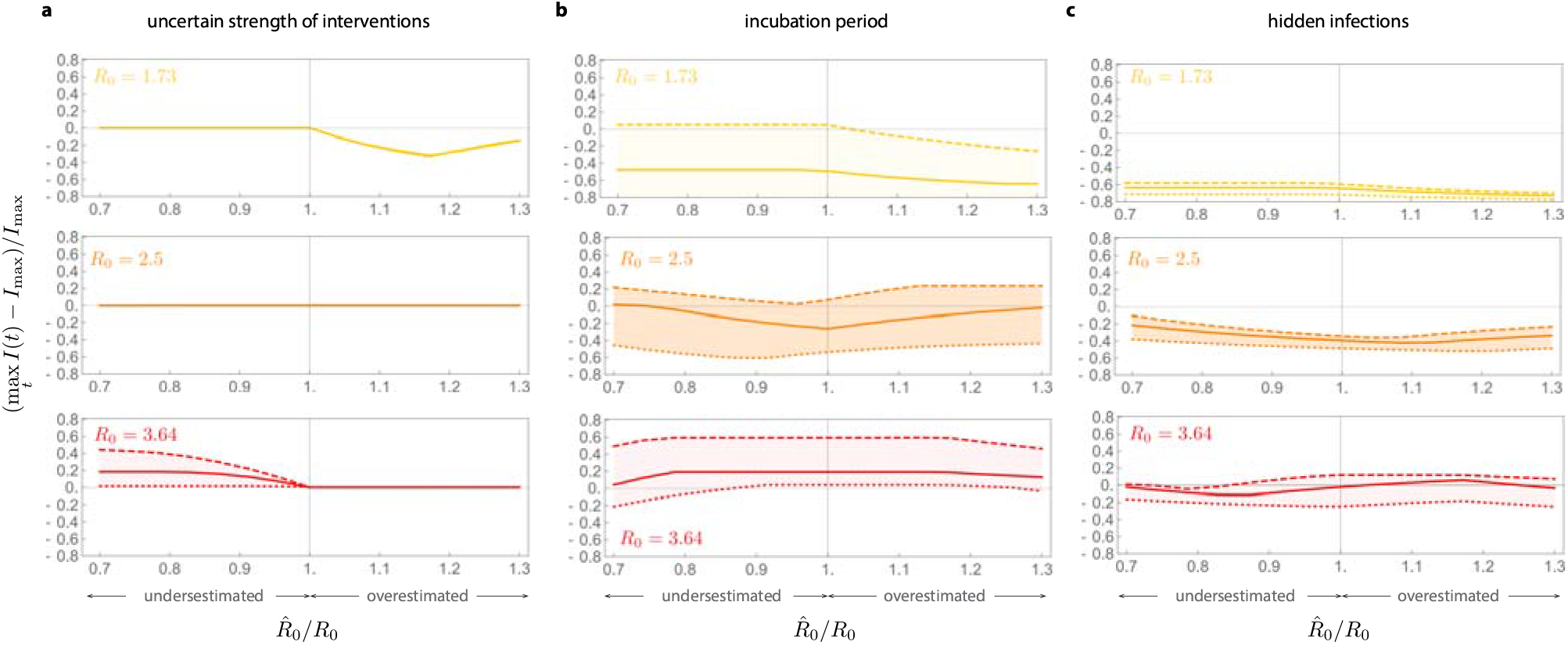
Optimal non-pharmaceutical interventions are robust. For all panels, the estimated parameters used for constructing the optimal NPIs are 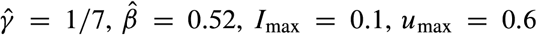. We consider a population of *N* = 8.855 x 10^6^ as in Mexico City, and the initial conditions *I*(0) = 1*/N* and *S*(0) = 1 − 1*/N*. If the models contain other state variables, they were initialized at zero. The optimal NPIs are constructed assuming 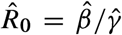, while the actual epidemic dynamics has a possibly Different *R*_0_ = *β/γ*. Panels shows results for outbreaks with three values of *R*_0_: low (yellow), medium (orange), and large (red). **a**. A SIR model where the reduction in the disease transmission by the NPIs is uncertain. We model this case replacing *u* by *ku* in the model equations. Panel shows the results for *k* = 1.1 (dotted), *k* = 1 (solid), and *k* = 0.9 (dashed). **b**. SEIR model where exposed individuals do not transmit the infection, with *λ* > 0 the incubation period. Panel shows the results for *λ* = 1/5 (dotted), *λ* = 1/7 (solid), and *λ* = 1/11 (dashed). **c**. A SEIIR model with *λ* = 1/7 and two classes of infected individuals (symptomatic and asymptomatic). Here, *p* ∈ [0, 1] is the proportion of exposed individuals that become asymptomatic. The vertical axis denotes the disease prevalence for symptomatic individuals. The panel shows the results for *p* = 0.55 (dotted), *p* = 0.7 (solid), and *p* = 0.8 (dashed).

For the final scenario, we consider an SEIIR model with an incubation period of 7 days and with a fraction *p* ∈ [0, 1] of infected individuals that are asymptomatic and thus remain hidden to the epidemic surveillance system. The goal is to maintain the prevalence of symptomatic individuals below *I*_max_, without knowing the fraction of asymptomatic individuals. This situation occurs during the COVID-19 pandemic, where between *p* = 0.55 and *p* = 0.8 of infections are asymptomatic [12]. For *p* < 0.7 and *R*_0_ < 3.64, the optimal NPIs maintain the disease prevalence of symptomatic individuals below or very close to *I*_max_ if the estimation error for *R*_0_ is below 30% (dotted and solid lines in Fig. 4c). An outbreak with low *R*_0_ produces a maximum disease prevalence of symptomatic individuals below *I*_max_, which may result in a larger effective duration of the interventions. Overall, these numerical results shows that the optimal NPIs are robust against a wide range of parameter uncertainty and unmodeled dynamics, provided that the estimation error in the outbreak’s basic reproduction number does not exceed 30%.

## Designing optimal NPIs for the COVID-19 pandemic

To explore the implications of our simple criterion for designing NPIs, we analyzed how 16 cities and regions implemented NPIs during the COVID-19 pandemic. For each region or city, we constructed *I*_max_ using the number of available intensive care beds, considering that a fraction of the infected individuals will require them (Supplementary Note S4). The *I*_max_ we obtain ranges from 2.87 x 10^−3^ for Lima (Peru) to 109.78 x 10^−3^ for Boston (US), reflecting the large heterogeneity of the available health services across the globe (Fig. 5a). With this information, we calculated the maximum feasible 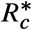 for each region using our design criterion of inequality (1). Since 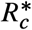 is a monotone function of *I*_max_, we find that 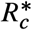 follows the same trend as *I*_max_ (Fig. 5b). The smallest 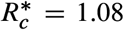 occurs for Lima and the largest 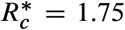 for Boston. Note that in both cases 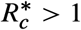. This result implies that, for the *R*_0_ of a region’s disease outbreak, NPIs policies must be implemented to guarantee that at least a reduction 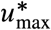 can be achieved such that 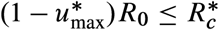.

**Figure 5:**
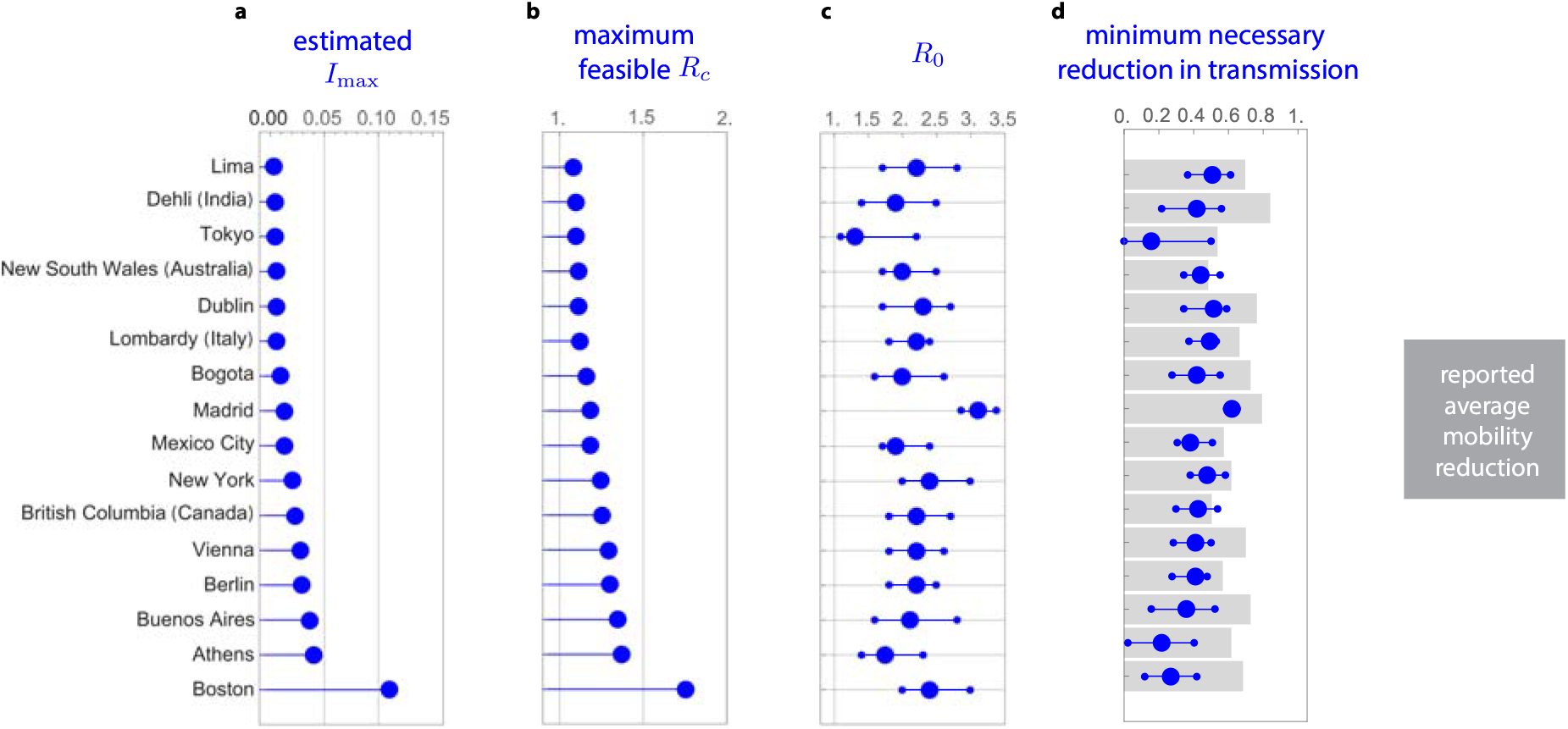
Minimum necessary reduction in disease transmission for NPIs in the COVID-19 pandemic. **a**. Calculated *I*_max_ according to the proportion of available intensive care beds in each region or city and the estimated fraction of infected individuals requiring intensive care. **b**. Maximum controlled reproduction number *R*_*c*_ that each region or city can handle according to its *I*_max_. Larger *I*_max_ allows a larger *R*_*c*_. **c**. Basic reproduction number *R*_0_ per region or city before interventions started. Median (blue big dot), and 95% confidence interval (smaller dots) are shown. **d**. Minimum *u*_max_ necessary for feasibility for each region or city (blue) according to the *R*_0_ of panel c. Grey bars denote the reported average mobility reduction in each region between March 19 and April 30.

Next, we investigated the *minimal* reduction 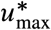 in transmission required to achieve those upper bounds for the COVID-19 pandemic. For this, we first collected information for the *R*_0_ in each region calculated at the start of the pandemic and when the NPIs were inactive (Supplementary Note S3). We find a median nominal *R*_0_ of 2.2, with Tokyo having the smallest one (*R*_0_ = 1.3) and Madrid having the largest one (*R*_0_ = 3.11), see Fig. 5c. From these values of *R*_0_, we calculated the minimal required reduction 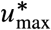 per region or city (blue in Fig. 5d). For the nominal *R*_0_’s per region or city, we find that a median reduction of 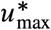 of 0.42 is necessary. However, this minimal necessary reduction is heterogeneous across regions. For example, Tokyo just requires 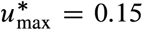 while Madrid requires 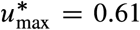. These two cities have the smallest and largest *R*_0_, respectively. If two cities have a comparable *R*_0_, then the city with large *I*_max_ ends requiring a smaller 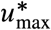 (e.g., Boston with 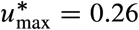 and Lima with 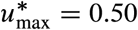).

To evaluate the feasibility of achieving the minimal reduction predicted by our analysis, we collected data for the average mobility reduction in each region during the NPIs in each region (grey in Fig. 5d and Supplementary Note S4). Considering this average mobility reduction as a proxy for the reduction in disease transmission, we find that all regions achieved a greater than necessary reduction. For example, Delhi attained a mobility reduction of 0.84, while the minimal necessary reduction in transmission according to our analysis is 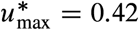. Other regions are in the boundary. For example, New South Wales attained a mobility reduction of 0.48, while the minimal necessary reduction in transmission was 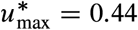. Overall, across regions, we find a median excess of 0.22 in the reduction of mobility compared to the minimal reduction in transmission 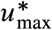 predicted by our analysis.

## Discussion and concluding remarks

Our choice of a simple SIR model was motivated by its epidemiological adequacy for the COVID-19 pandemic and its low dimensionality. The mathematical tractability of the SIR model gives us a complete understanding of the optimal NPIs to apply at any epidemic state. The feedback form *u*^∗^(*S, I*) of the optimal intervention reflects such understanding, prescribing the optimal action to perform if the epidemic is at state (*S, I*). This feedback strategy should be contrasted to most other studies applying optimal control to epidemic outbreaks, where the optimal intervention is written as an open-loop function of time *u*^∗^(*t*) [13–16] (see Supplementary Note S4 for details about how our work is related to existing optimal control studies). The open-loop intervention gives the optimal action at any time for a particular initial state. However, it does not tell us what the optimal action is if the epidemic is not in the exact state predicted by the model. Understanding the optimal action to perform at any state has the crucial advantage of allowing us to apply this knowledge to any model, and therefore to reality. Indeed, feedback gives control strategies the required robustness to work on real processes [17, 18], and we numerically confirm that the optimal NPIs we derived have such robustness. Future work could analyze the robustness of the optimal intervention when the state of the epidemic is not entirely known. For example, this case may happen when significative delays exist in reporting new infections, or when tests for identifying infected individuals are limited.

The optimal intervention resulting from our analysis can take a continuum of values that may be infeasible to implement in practice. We can use an averaging approach to circumvent this problem. Namely, consider a time window of *T* days (e.g., a week). Suppose that the average reduction prescribed by the optimal intervention over a certain window is *ū*^∗^. We can realize this reduction on average by combining *d* = *T ū*^∗^/*u*_max_ days of maximum reduction with (*T* − *d*) days without intervention. This approach yields an intervention similar to Karin et al. [19], with the difference that the periods of intervention and activity are optimally balanced.

Our criterion to design optimal NPIs for mitigating epidemic outbreaks is obtained by characterizing the necessary and sufficient conditions for the existence of solutions to an optimal control problem. Specifically, the low-dimensionality of the SIR model allowed us to apply Green’s Theorem to compare the cost of any two interventions analytically (Supplementary Note S1.4). In this sense, the method we use to derive the optimal NPIs is closer to our previous work on optimal control for bioreactors [20]. In general, deriving such complete characterization of optimal control problems is challenging because it involves solving an infinite-dimensional optimization [21]. Indeed, computational methods cannot produce such a characterization [22], and established analytical methods like Pontryagin’s Maximum Principle only yields necessary conditions for optimality [21]. We note that there are several studies applying these and other similar methods to the SIR model [23, 24], in particular during the COVID-19 pandemic [11, 25–28]. Our results could guide a complete characterization of optimal NPIs for more detailed epidemic models or more detailed optimization objectives, but this is likely very challenging.

We will inevitably face new epidemics where non-pharmaceutical interventions are the only option to control infections. Rather counter-intuitively, we find that for “ending” an epidemic outbreak as fast as possible using NPIs it is not always optimal to apply the maximum intervention. This observation illustrates the need for developing a better scientific understanding that can inform the design of optimal non-pharmaceutical interventions and plan the required health services capacity.

### BOX 1.

**Optimal NPIs for the Susceptible-Infected-Removed (SIR) model**

The SIR model with interventions *u*(*t*) ∈ [0, *u*_max_] reducing disease transmission takes the form

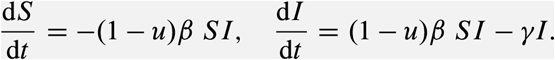

Here, *S*(*t*) and *I*(*t*) are the proportion of the population that is susceptible or infected at time *t* ≥ 0, respectively. We denote by (*S*_0_; *I*_0_) the initial state at *t* = 0. The parameters of the SIR model are the (effective) *contact rate β* ≥ 0, and the mean *residence time* of infected individuals *γ* ≥ 0 (in units of day^−1^). By assuming *S*_0_ ≈ 1, these two parameters yield the *basic reproduction number R*_0_ = *β/γ*.

We are interested in reaching the *safe zone*

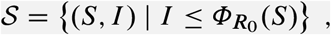

where

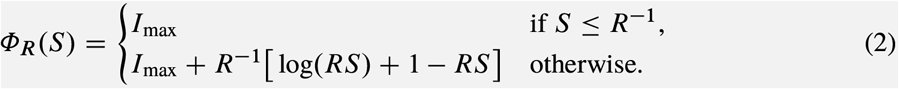

The safe zone is the largest set with the following property: If, for any given time *t*_1_, the state (*S*_1_; *I*_1_) belongs to *𝒮*, we can set *u* = 0 henceforth and still have *I*(*t*) ≤ *I*_max_ for all *t* ≥ *t*_1_. That is, when *𝒮* is reached, we can terminate the intervention with the assurance that a possible rebound in the disease prevalence will not exceed *I*_max_.

Our goal is to steer an arbitrary initial state (*S*_0_; *I*_0_) to the safe zone *𝒮* in minimal time without violating the constraint *I*(*t*) ≤ *I*_max_. We say that an intervention achieving this goal is an *optimal intervention*. In Supplementary Note S1, we prove that the existence of an optimal intervention is characterized by the *separating curve* 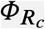 as follows:

1. An optimal intervention exists if and only if the initial state (*S*_0_; *I*_0_) lies below this separating curve (i.e., 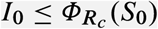). Above, *R*_*c*_ := (1 − *u*_max_)*R*_0_ is the *controlled reproduction number*. Moreover:
2. If it exists, the optimal intervention *u*^∗^ at the state (*S, I*) is

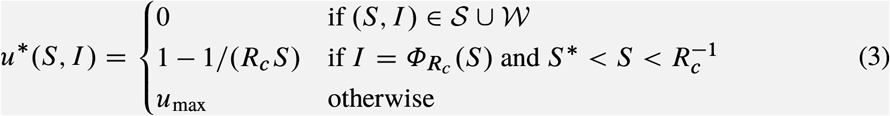

with

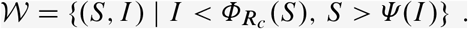

Above, the curve *S* = *Ψ* (*I*) is defined in Supplementary Note S1, while *S* ^∗^ denotes the intersection of *S* = *Ψ* (*I*) and 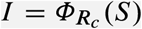.

## Data Availability

Data is included in Supplementary Note 3

## Supplementary Notes

### S1. Characterization of the optimal intervention in the Susceptible-Infected-Removed model

The model is given by

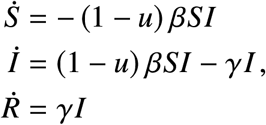

where the parameters *β* > 0, *γ* > 0 are assumed constant. Since the total population *N* = *S* + *I* + *R* remains constant all the time, the model can be reduced to that of a second order system using only the states (*S, I*). The maximal (acceptable) value of *I* is *I*_max_ and the maximal achievable value of the control is *u*_max_. So the state has to belong to the following feasible sets

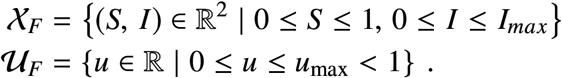

Sometimes it will be useful to write the Differential equation in a compact form as

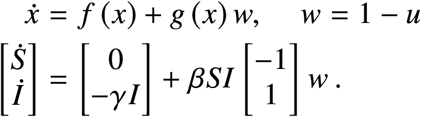

The trajectory starting at the initial point *x*_0_ = (*S*_0_, *I*_0_) and subject to the control *u* : ℝ → 𝒰_*F*_ is denoted by *ϕ* (*t, x*_0_, *u* (·)).

Let us define the function

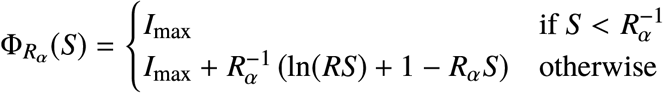

with *R*_*α*_ ∈ {*R*_*c*_, *R*_0_}. The optimal control problem consists in finding the control strategy *u* such that, starting from the initial point (*S*_0_, *I*_0_), the target set

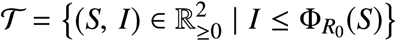

is reached in the minimal time with the state restriction *I* (*t*) ≤ *I*_max_ satisfied for all time. Note that this set is positively invariant without control (*u* = 0), and that every trajectory that starts in this set satisfies the restriction *I*(*t*) ≤ *I*_max_ for all *t* ≥ 0 (see Fig. S1).

Now let us define the reachable set for an initial state *x*_0_ as the set of points that can be reached from the initial point *x*_0_ with feasible control, i.e.,

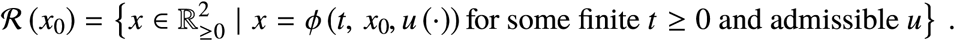

Also, we define the controllable set of the target set 𝒯 as the set of points from which some point in the target 𝒯 can be reached with a feasible control, i.e.,

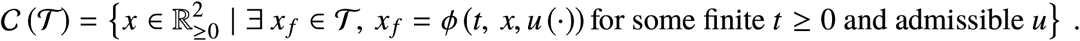

The set 𝒞 (𝒯) can be equivalently described as ℛ (𝒯) for the system

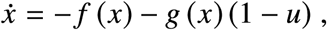

**Supplementary Figure S1.**
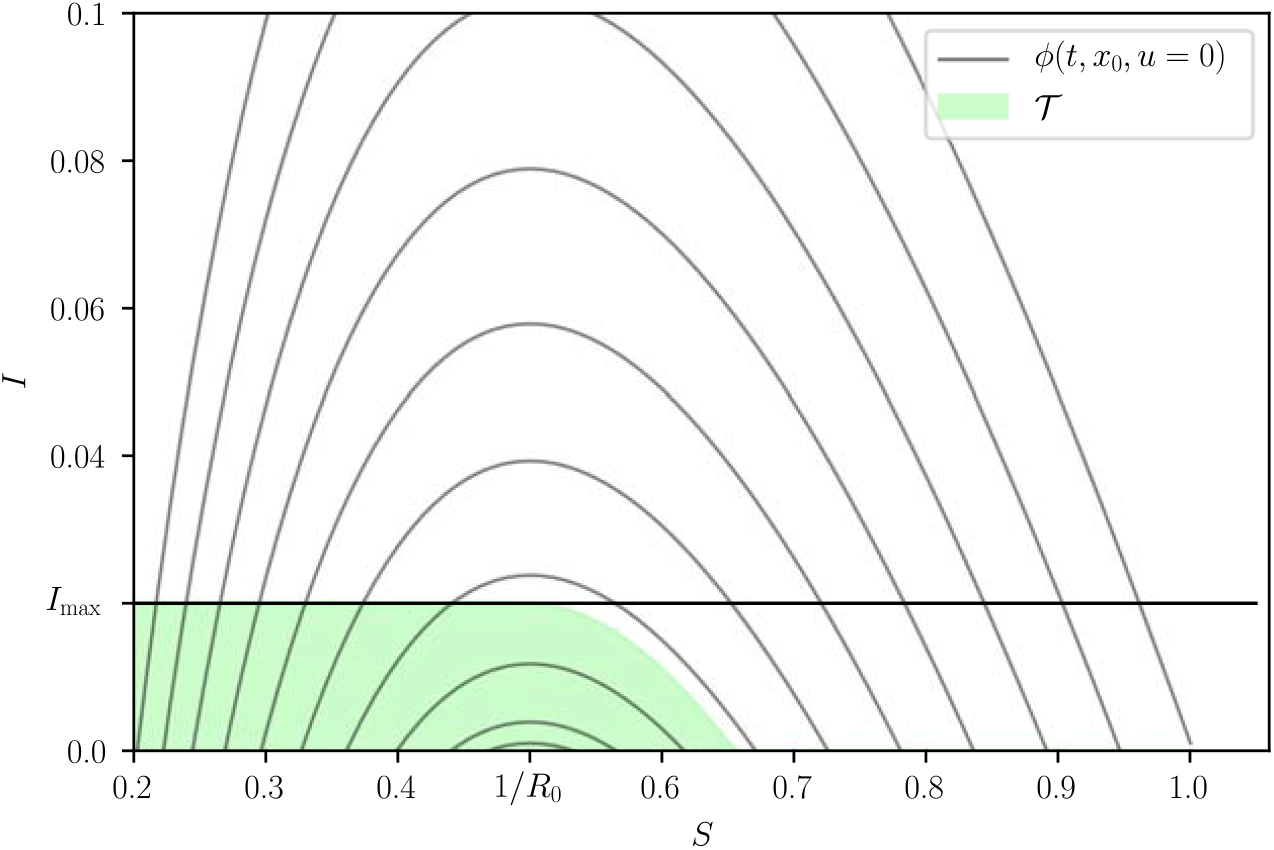
The set 𝒯 is the largest positive invariant set satisfying *I* ≤ *I*_max_. The figure was generated with *R*_0_ = 2, *R*_*c*_ = 1.18 and *I*_max_ = 0.02.

i.e., the set of points that can be reached from the set 𝒯 for the dynamics with backward time. Now, the optimal control problem has a solution if and only if

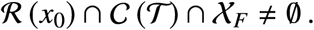

Since the points of the form (*S, I*) = (*S*, 0) are equilibria for every control value, ℛ ((*S*, 0)) = (*S*, 0), we exclude them from the initial conditions for which there is a solution (except if the equilibrium is already in the target set). Now, since 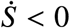 for *S* > 0, *I* > 0,

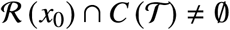

for every initial condition (except for initial conditions of the form (*S*, 0)). It is obvious that, for the problem to be feasible, the initial state has to be in the feasible set 𝒳_*F*_, i.e.,

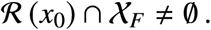

#### S1.1 Calculation of the orbits

Although it does not seem to be possible to find the trajectories of the system explicitly, it is easy to find its orbits. For this we write (we exclude the points for which *I* = 0 since they are equilibria)

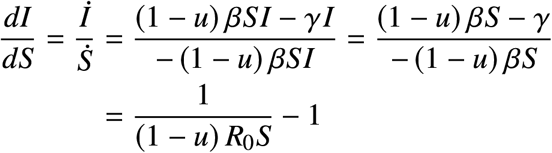

which is a separable Differential equation (DE). Assuming that *u* is constant and integrating, we obtain

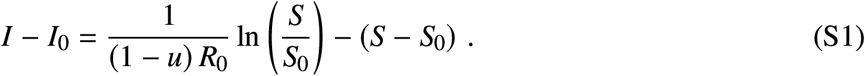

An interesting rewriting of (S1) is

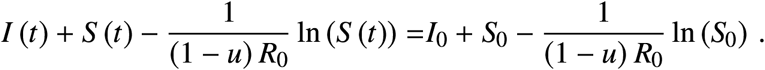

This means that the quantity 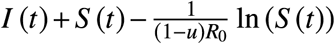 remains constant along the trajectory. Note that this constant depends on the control value used. The above equation is well-known for the SIR model (see, e.g., [1]).

Given an initial condition (*S*_0_, *I*_0_) this expression gives, for any 0 < *S* < *S*_0_ the (unique) value of *I* that is reached in future time^1^. Thus there exists a function *I* (*S*; (*S*_0_, *I*_0_)) that gives the value of *I* as a function of *S* and the initial condition. Moreover, from the first equation in the DE we obtain

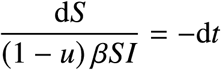

and, if we take the expression *I* (*S*; (*S*_0_, *I*_0_)), we obtain a separable DE that can be integrated,

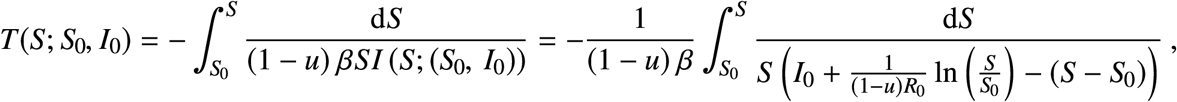

and that gives the time to reach the point (*S, I* (*S*)) from the initial point (*S*_0_, *I*_0_) with the (constant) control *u*. Although it does not seem possible to give an explicit expression for this integral, it is clear that *S* parametrizes uniquely the solutions (since it is monotone).

#### S1.2 The number of infected people

If we apply a constant control 0 ≤ *u* ≤ *u*_max_ the infection will eventually die out, i.e., the value *I* (∞) = 0 will be reached asymptotically (otherwise *R* (*t*) would continue growing, which is impossible). We can therefore compute *S* (∞) implicitly from (S1) as

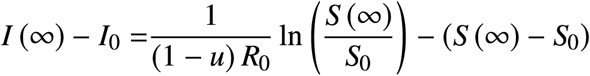

or, equivalently, as

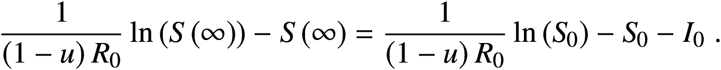

Note that the final value of *S* depends on the initial values, but also on the control used.

If we assume that the model is normalized, and the initial value is *S*_0_ = 1 and *I*_0_ ≈ 0, then

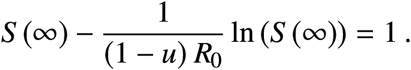

Note that, if *u* → 1^−^, then *S* (∞) → 1^−^. So, the larger the value of *u*, the larger the value of *S* (∞).

#### S1.3 Reachable set from (*S*_0_, *I*_0_)

At each point in the state space, the directions in which the vector field points for Different values of the control are given by *F*_*u*_ (*x*) = *f* (*x*) + g (*x*) (1 − *u*). The extreme values are given by *F*_0_ (*x*) = *f* (*x*) + g (*x*) and 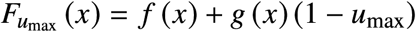,

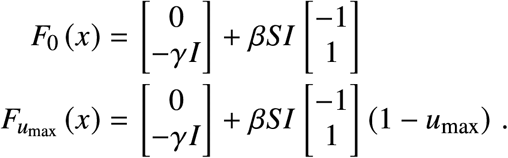

**Supplementary Figure S2.**
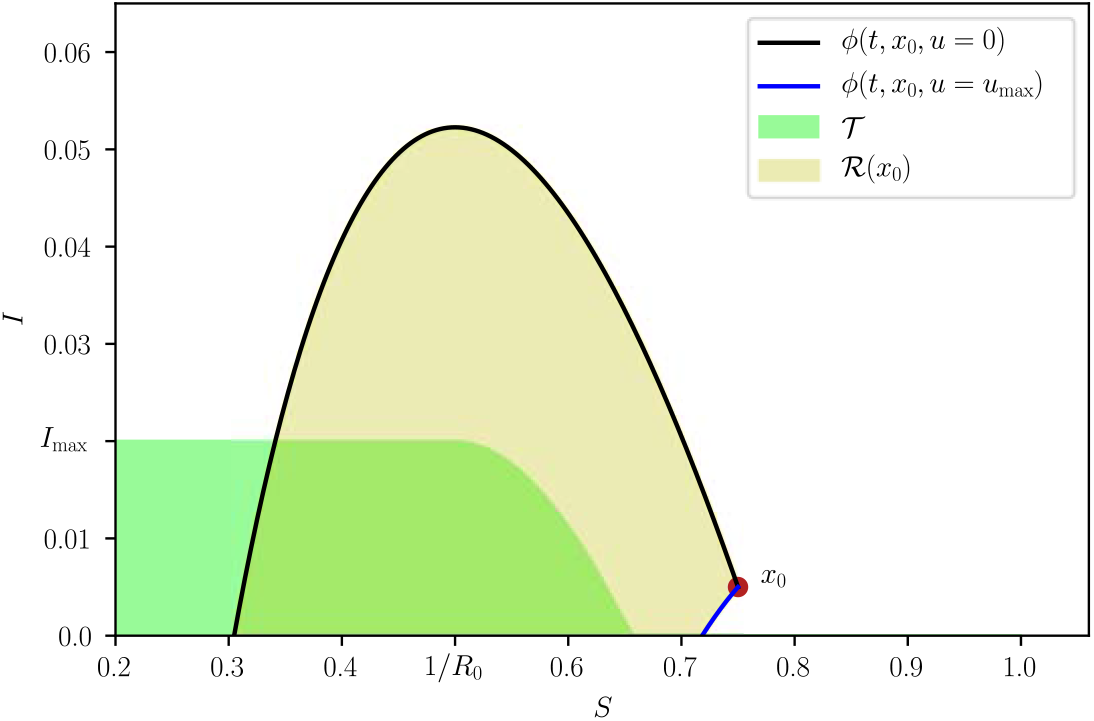
Phase Plane with maximal and minimal orbits bounding the reachable set ℛ (*x*_0_). Max corresponds to the trajectory *ϕ* (*t, x*_0_, *u* = 0) while Min to *ϕ* (*t, x*_0_, *u* = *u*_max_). The figure was generated with *R*_0_ = 2, *R*_c_ = 1.18 and *I*_max_ = 0.02.

In the phase plane (*S, I*) both point to the “left”, since the first component (in the direction of *S*) is always negative (recall that *SI* > 0). Since for the second components of the vector fields we have

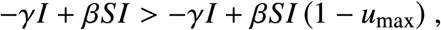

it follows that *F*_0_ is “above” 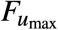. Therefore, the reachable set ℛ (*x*_0_) is bounded by the two trajectories *ϕ* (*t, x*_0_, *u* = 0) and *ϕ* (*t, x*_0_, *u*_max_), see Fig. S2. These two bounding orbits can be easily calculated using Eq. (S1).

#### S1.4 Comparing the cost of two Different trajectories

In order to be able to find the orbit (trajectory) solving the optimal control problem, it is necessary to be able to compare the cost of two Different trajectories that start at the same initial point and end at the same final point. Consider two orbits *ω*_*i*_ (*x*_0_, *x*_*f*_, *u*_*i*_), *i* = 1, 2, joining the (same) points *x*_0_ and *x*_*f*_ using two Different control actions, *u*_1_ and *u*_2_, respectively. The cost (*i*.*e*. time) going through *ω*_*i*_ is

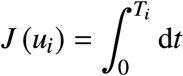

along the trajectory. Given two such orbits, we want to compare both costs. This can be done, for example, by subtracting them, i.e., if

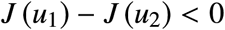

then the cost of *ω*_1_ is lower than that of *ω*_2_.

The cost *J* (*u*_*i*_) can be calculated as a line integral along the trajectory. We can see this in the following manner. Calculate

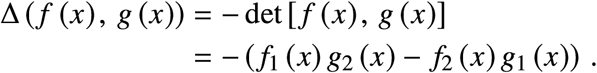

Now, by properties of the determinant this is also the same as

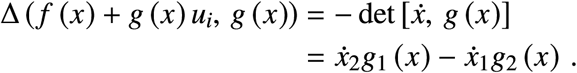

Therefore,

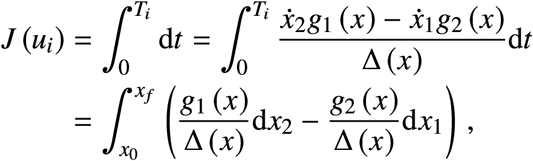

which is a line integral along the orbit *ω*_*i*_. Since the two paths have the same initial and final points, they form a closed curve, and calculating the line integral along the closed curve followed in the counterclockwise direction we obtain the difference of the costs, i.e.

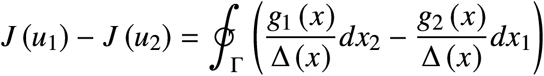

where Γ is the closed path of the two orbits followed in the counterclockwise direction. For this we have to assume that: (1) the two paths (orbits) do not intersect at any points except the initial and final ones, and (ii) that Δ ≠ 0.

Using Green’s theorem, the line integral can be calculated using a surface integral:

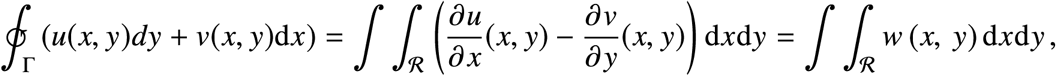

where ℛ is the region enclosed by the closed curve Γ. For our problem this becomes

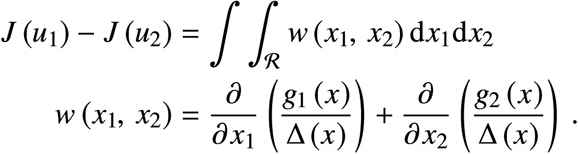

In our case,

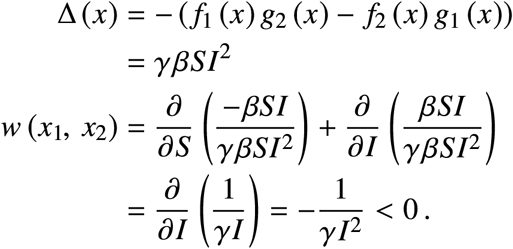

We see that *w* < 0 everywhere, and therefore the integral is always negative, implying that the “upper” orbit has a lower cost than the “lower” orbit (in the closed path traversed in the counterclockwise direction). This observation allows us to find the optimal orbit by comparing it with others.

#### S1.5 Optimal orbits

From the previous results, the “upper” trajectory is the one with no control (*u* = 0) and, in terms of the cost alone, this trajectory is better than any other one joining the same two points. However, such control may be inadmissible, since the corresponding *I* can go over *I*_max_ at some periods of time.

The computation of the optimal control can be approached in two ways:

- Fix the initial condition *x*_0_, find its optimal orbit and then its associated optimal control.
- Study the optimal control problem for all possible initial conditions.

Although the second approach is obviously better, it is more difficult, so we will start with the first approach. In fact, both approaches should lead to the same conclusions.

Now we can divide the study of the optimal orbit in several cases.

##### S1.5.1 Unfeasible trajectories

This is the case if *I*_0_ > *I*_max_.

##### S1.5.2 Trivial trajectories

This is the case in which 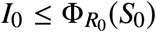. That is, the case in which we start in the target set.

##### S1.5.3 Bang-bang trajectories

If *x*_0_ ∉ 𝒯, it is necessary to apply some control to maintain *I* below the maximal value *I*_max_. Moreover, admissible trajectories necessarily cross the boundary of 𝒯 at *S* ≥ 1/*R*_0_, that is, they enter 𝒯 at

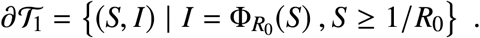

In order to find the optimal control that steers an initial state *x*_0_ to *x*_*f*_ ∈ ∂𝒯_1_, consider the change of coordinates

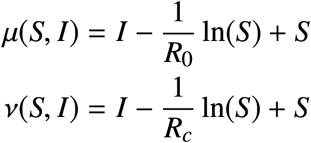

with inverse

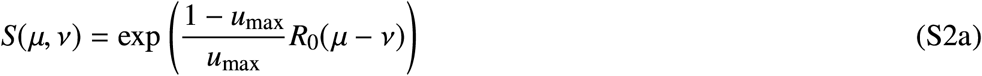

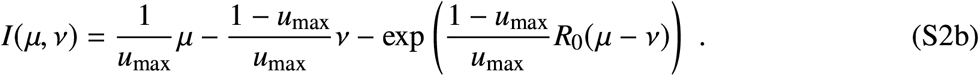

Note that, given *I* = 0, we can uniquely map *μ* to *S* ≥ 1/*R*_0_, which we will denote by *S* = *Ŝ*_*μ*_(*μ*). Likewise, we can uniquely map *ν* to *S* ≥ 1/*R*_*c*_, and we will denote it by *S* = *Ŝ*_*ν*_ (*ν*).

In the new coordinates, the dynamic equations are

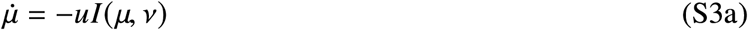

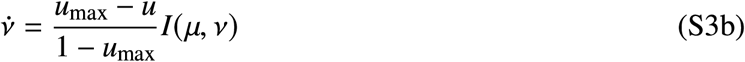

Note that *μ* = const is an orbit when *u* = 0 and *ν* = const is an orbit when *u* = *u*_max_.

In (*μ, ν*)-coordinates, the entry point at 𝒯 is the segment

**Supplementary Figure S3.**
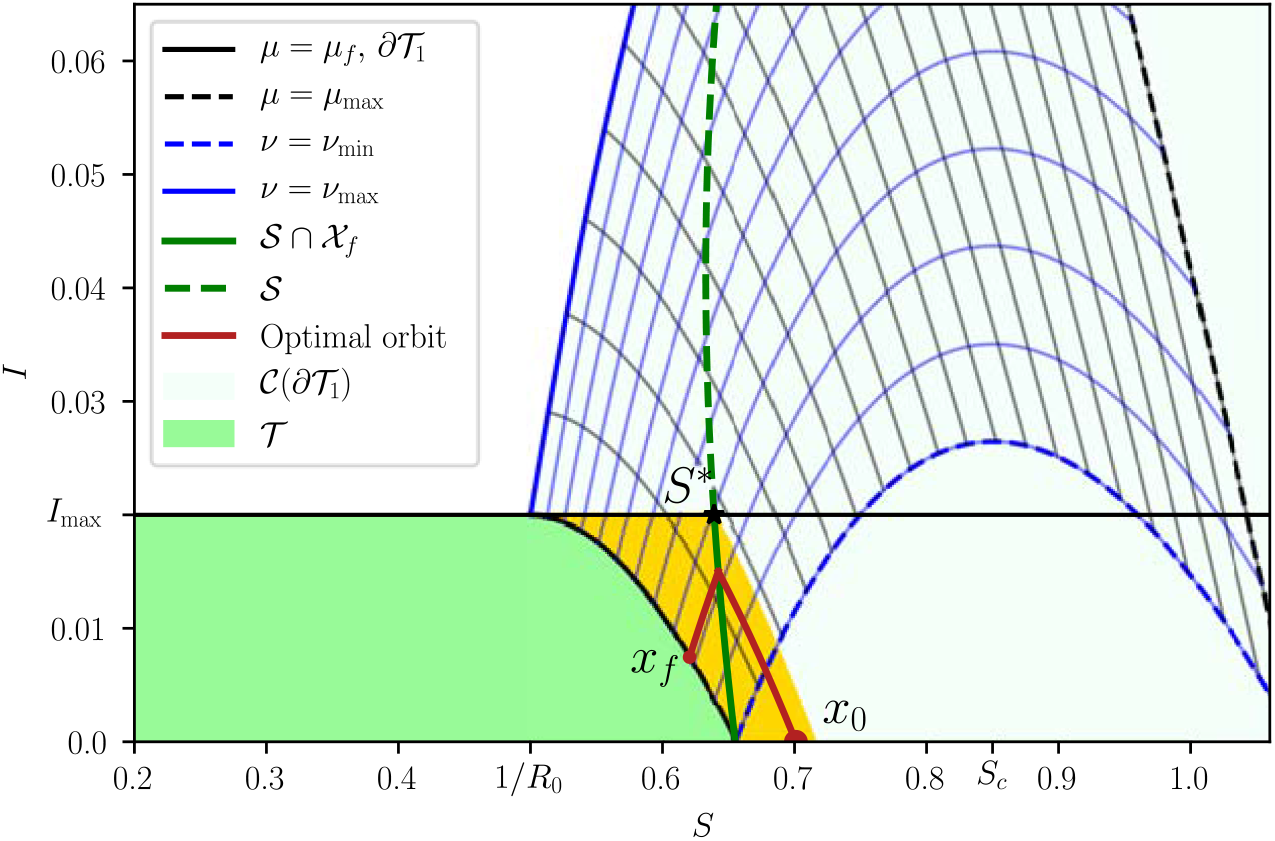
An optimal bang-bang trajectory that initiates at *x*_0_ with *u* = 0 (red). When the state touches the curve 𝒮 (green), the control is set to *u* = *u*_max_ until the state reaches *x* _f_ ∈ ∂𝒯_1_. The region containing all bang and bang-bang trajectories is depicted in yellow. The figure was generated with *R*_0_ = 2, *R*_c_ = 1.18 and *I*_max_ = 0.02.

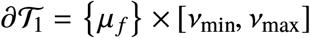

with

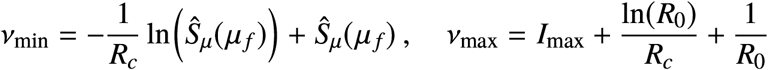

and

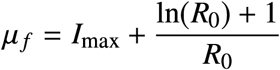

(see Fig. S3).

It follows from Sec. S1.4 that the fastest orbit joining an initial state (*μ*_0_, *ν*_0_) ∈ 𝒞(∂𝒯_1_) and a final state (*μ*_*f*_, *ν*_*f*_) ∈ ∂𝒯_1_ is the concatenation of a first piece connecting (*ν*_0_, *μ*_0_) and (*ν*_*f*_, *μ*_0_) with *u* = 0 and a second piece connecting (*ν*_*f*_, *μ*_0_) and (*ν*_*f*_, *μ*_*f*_) with *u* = *u*_max_. That is, the control is bang-bang. It is easy to verify that this control yields the fastest trajectory, as any other trajectory joining (*μ*_0_, *ν*_0_) and (*μ*_*f*_, *ν*_*f*_) is below this one. The transition times can be computed using (S3) as

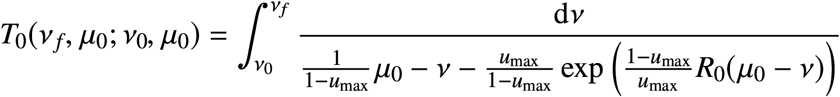

and

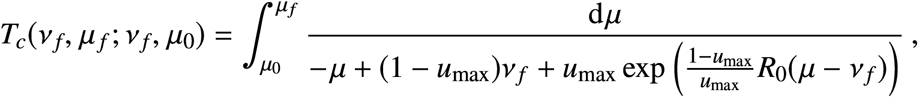

so that the total time is *T* (*ν*_*f*_, *μ*_*f*_; *ν*_0_, *μ*_0_) = *T*_0_(*ν*_*f*_, *μ*_0_; *ν*_0_, *μ*_0_) + *T*_*c*_(*ν*_*f*_, *μ*_*f*_; *ν*_*f*_, *μ*_0_).

Note that *μ*_*f*_ is fixed, but *ν*_*f*_ ∈ [*ν*_min_, *ν*_max_] is free. We will now find the closest entry point by minimizing *T* over *ν*_*f*_. Set

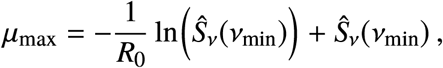

**Supplementary Figure S4.**
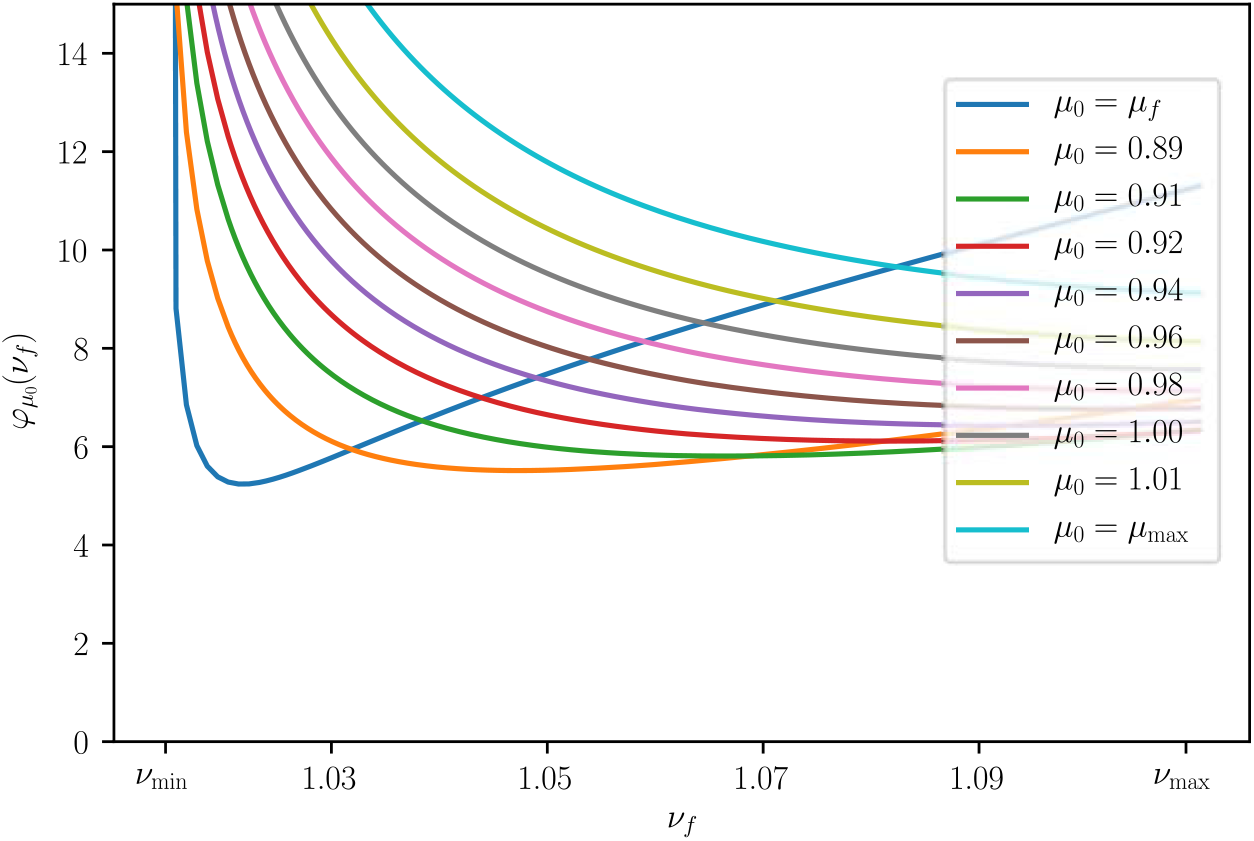
The minimal time *φ*_*μ*_ (*ν*_f_) it takes an initial state (*ν*_min_, *μ*_0_) to reach the target point (*ν*_f_, *μ*_f_). The curves were generated with *R*_0_ = 2 and *R*_c_ = 1.18. Note that the global minimum is well defined (it is unique).

fix *μ*_0_ ∈ [*μ*_*f*_, *μ*_max_] and define the map

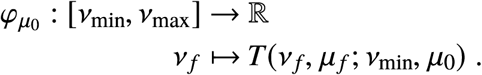

**Assumption 1**. *Global minima of* 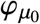 *are unique*.

By Weierstrass Theorem, global minima of 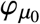 always exist. The assumption excludes the highly degenerate case in which the global minimum could occur for more than one value of *ν*_*f*_. Figure S4 shows plots of 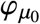 for various values of *μ*_0_ using the parameters *R*_0_ = 2 and *R*_*c*_ = 1.18. Note that the global minimum is unique (indeed, for large values of 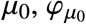 is convex). We now define the function

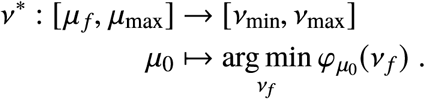

This function defines a switching curve parameterized by *μ*_0_. In the original coordinates (S2), the switching curve takes the form

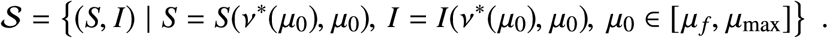

Let *Ī* = max_(*S,I*)∈𝒮_. To simplify the exposition, we introduce Ψ : [0, *Ī*] → [0, 1], defined implicitly by

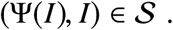

We will parameterize 𝒮 using *I*,

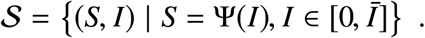

The trajectories that reach 𝒮 above *I*_max_ are of course unfeasible, so the class of optimal bang-bang trajectories are only those that pass through 𝒮∩𝒳_*F*_ (see the yellow region in Fig. S3). For future reference, we will denote by *S*^∗^ the *S* coordinate at which 𝒮 intersects the line *I* = *I*_max_ and by *x*_1_ the point (*S*^∗^, *I*_max_).

Summarizing, there are two possible situations:

1. **Bang**. If *x*_0_ belongs to the region delimited by ∂𝒯_1_, *I* = *I*_max_ and 𝒮, then the optimal control strategy is simply

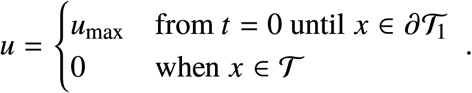
2. **Bang-Bang**. When *x*_0_ belongs to the region delimited by 𝒮, *I* = *I*_max_ and the orbit *ϕ*(−*t, x*_1_, *u*_max_), then the optimal control strategy is

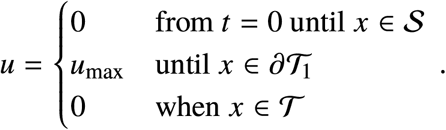

##### S1.5.4 Trajectories containing a singular arc

Let us define an initial point *x*_0_ = (*S*_0_, *I*_0_) and the point *x*_1_ = (*S*^∗^, *I*_max_). We are interested in four trajectories (or orbits):

1. *ϕ* (*t, x*_0_, *u* = 0), the trajectory without control starting at *x*_0_. It will be useful to calculate the value 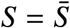 at which the orbit (first) touches *I*_max_. For this we solve (use (S1))

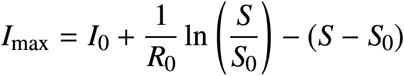

for *S* and obtain two solutions: *S*_1_, *S*_2_. Define 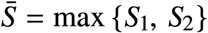 as the largest. Now we calculate the values of *S* ≤ *S*_*c*_ for which it is possible to achieve *İ* ≤ 0 (that is, that it is possible to stop the growth of *I*). This value can be calculated from

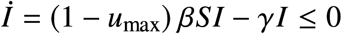

and gives

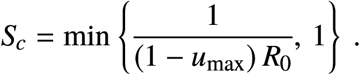

We “saturate” the value of *S*_*c*_ because *S*_*c*_ > 1 is not empidemiologically relevant. The control required to achieve the condition *I* = *I*_max_ is the “singular” control

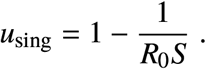

Note that, if *S* > *S*_*c*_ at *I* = *I*_max_, it is no longer possible to keep *I* at *I*_max_ because *İ* > 0. If 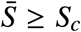, then the optimal control is **bang-singular arc-bang**,

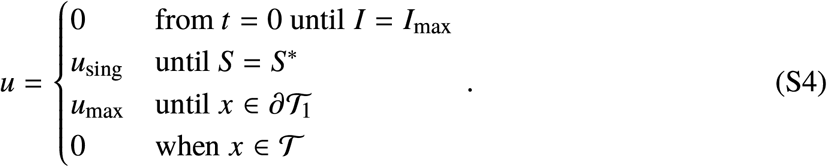

This case is depicted in Fig. S5 for the parameters *R*_0_ = 2, *R*_*c*_ = 1.18 and *I*_max_ = 0.02. If *S*_*c*_ = 1, then 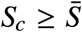 holds trivially and the optimal strategy is again (S4).
2. *ϕ* (*t, x*_0_, *u*_max_), the trajectory with maximal control starting at *x*_0_.
3. *ϕ* (−*t, x*_1_, *u* = 0), the trajectory without control that passes through *x*_1_. If *x*_0_ is at the left of this trajectory, the optimal orbit is bang-bang, as shown in the previous section. Optimal trajectories starting at the right have singular arcs.
4. *ϕ* (−*t, x*_1_, *u*^∗^), the trajectory with control

**Supplementary Figure S5.**
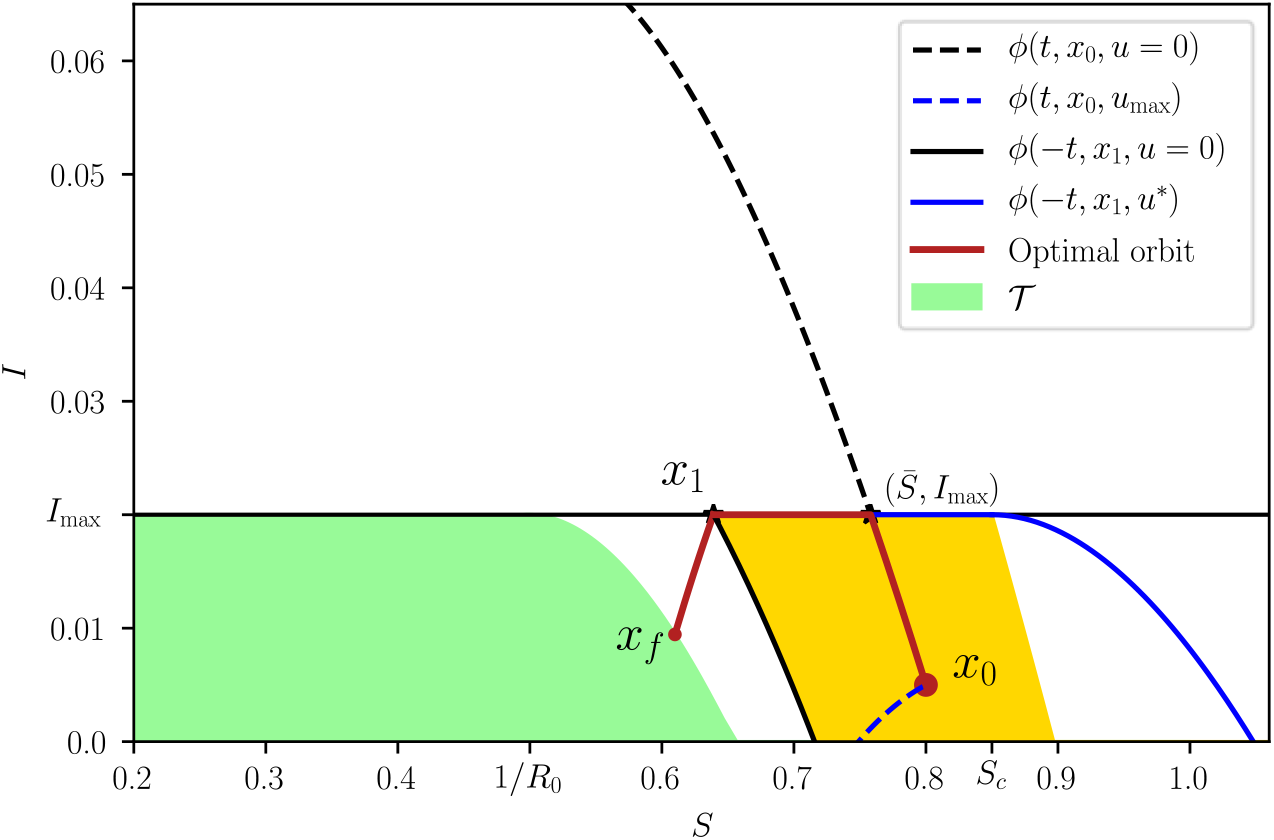
An optimal trajectory that initiates at *x*_0_ with *u* = 0 (red) and such that 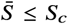. When the state touches the curve *I* = *I*_max_, the control is set to *u* = *u*_sing_ until the state reaches *x*_1_. The control is finally switched to *u* = *u*_max_ until the state reaches the target set. The region containing all bang-singular arc-bang trajectories is depicted in yellow. The figure was generated with *R*_0_ = 2, *R*_c_ = 1.18 and *I*_max_ = 0.02.

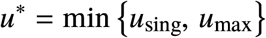

that passes through *x*_1_.

The control *u*^∗^ is such that this trajectory does not violate the restriction *I* ≤ *I*_max_. For values of *S* ≥ *S*_*c*_, it is equal to *u*_max_, and for *S* ≤ *S*_*c*_ it is the control for the singular arc, i.e., it maintains *I* = *I*_max_ until *x*_*f*_ is reached.

When 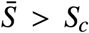 then it is necessary to start with the control strategy before reaching the maximal value of *I* = *I*_max_. Otherwise, this limit will be surpassed. However, this is only feasible if, moving backwards from the point (*S*_*c*_, *I*_max_) with the maximal control *u*_max_ it is possible to reach a point (*S*_0_, *I*_*c*_) such that *I*_*c*_ ≥ *I*_0_. The value of *I*_*c*_ can be calculated from (S1),

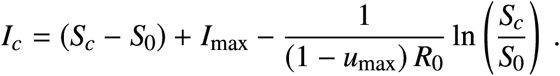

If *I*_*c*_ = *I*_0_, the optimal control is

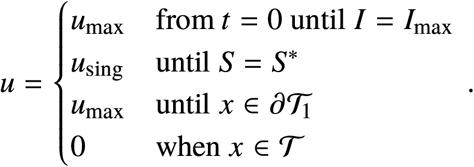

When *I*_*c*_ > *I*_0_, the control is **bang-bang-singular arc-bang**,

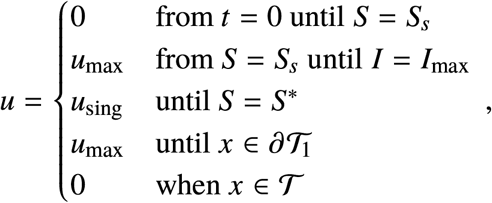

where (*S*_*s*_, *I*_*s*_) is a switching point. It is characterized as follows: the trajectory *ϕ* (*t, x*_0_, *u* = 0) intersects the trajectory *ϕ* (−*t, x*_1_, *u*_max_) at (*S*_*s*_. *I*_*s*_). Such point can be calculated from (S1) as

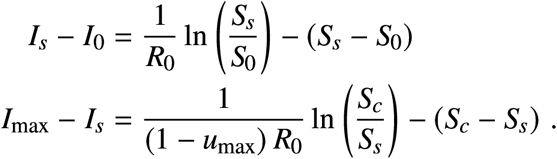

By substituting the first into the second we get

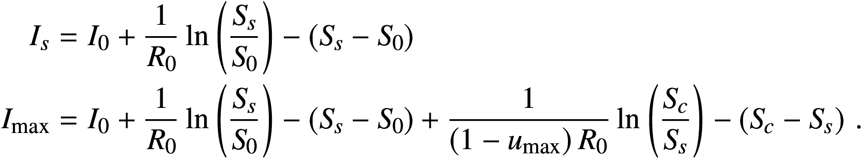

Solving for *S*_*s*_ in the second we arrive at

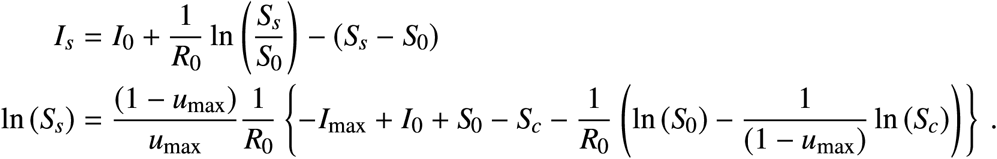

This case is depicted in Fig. S6 again for the parameters *R*_0_ = 2, *R*_*c*_ = 1.18 and *I*_max_ = 0.02.

If *I*_*c*_ < *I*_0_, then it is not possible to solve the optimal problem, since any strategy will surpass the maximal value *I*_max_. This is the case if, e.g., *u*_max_ is reduced and *R*_*c*_ increases to 1.27 (see Fig. S7).

#### S1.6 A feedback control strategy

The previous “open loop” strategy can be implemented as a state feedback control. This strategy is rather simple, since there are basically only two switching curves: *ϕ* (−*t, x*_1_, *u*^∗^) and 𝒮. Another switch takes place when the target region has been attained and the control is switched off, but this happens in a “natural” manner.

The switching curve *ϕ* (−*t, x*_1_, *u*^∗^) can be written as

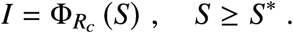

We can further define the “waiting” set

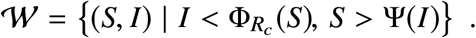

**Supplementary Figure S6.**
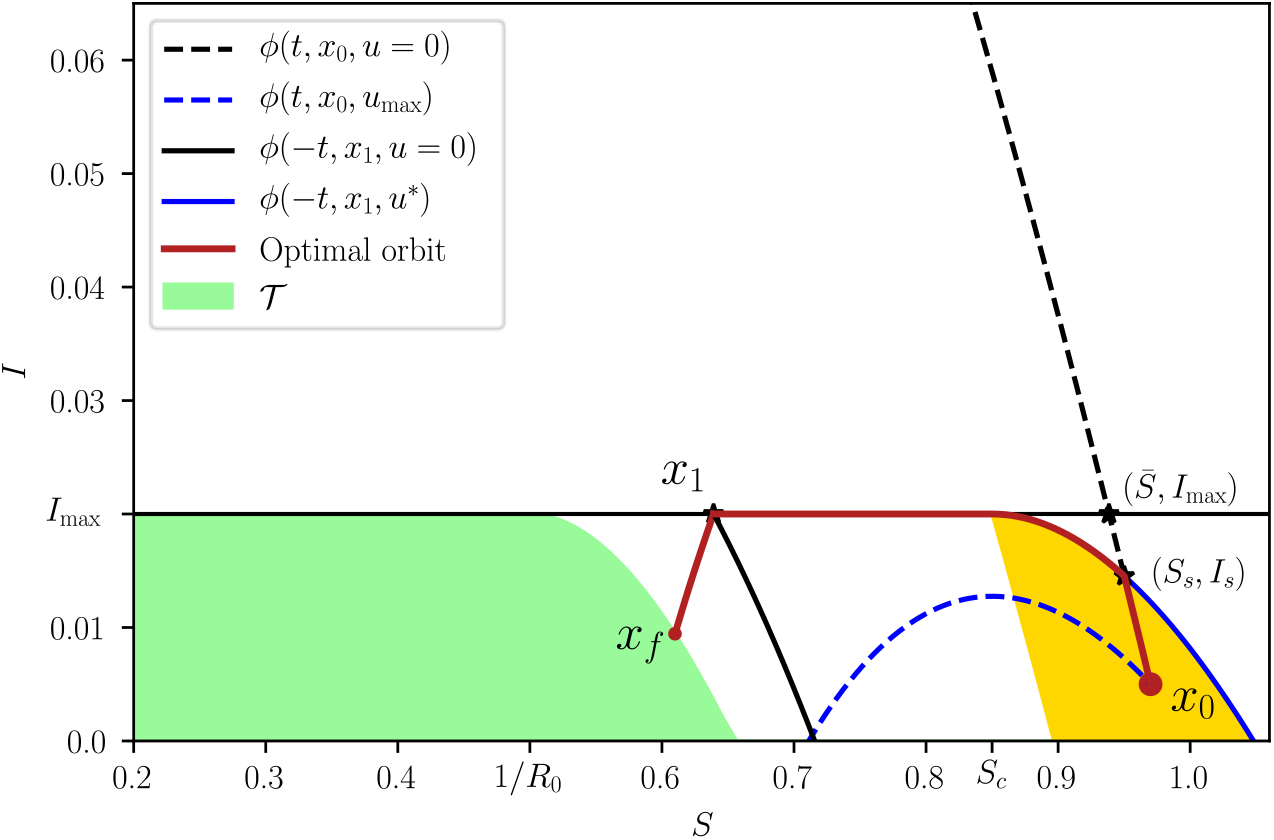
An optimal trajectory that initiates at *x*_0_ with *u* = 0 (red) and such that 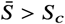. When the state arrives at (*S*_*s*_, *I*_*s*_), the control is set to *u* = *u*_max_. When the state touches the curve *I* = *I*_max_, the control is set to *u* = *u*_sing_ until the state reaches *x*_1_. The control is finally switched to *u* = *u*_max_ until the state reaches the target set. The region containing all bang-bang-singular arc-bang trajectories is depicted in yellow. The figure was generated with *R*_0_ = 2, *R*_*c*_ = 1.18 and *I*_max_ = 0.02.

**Supplementary Figure S7.**
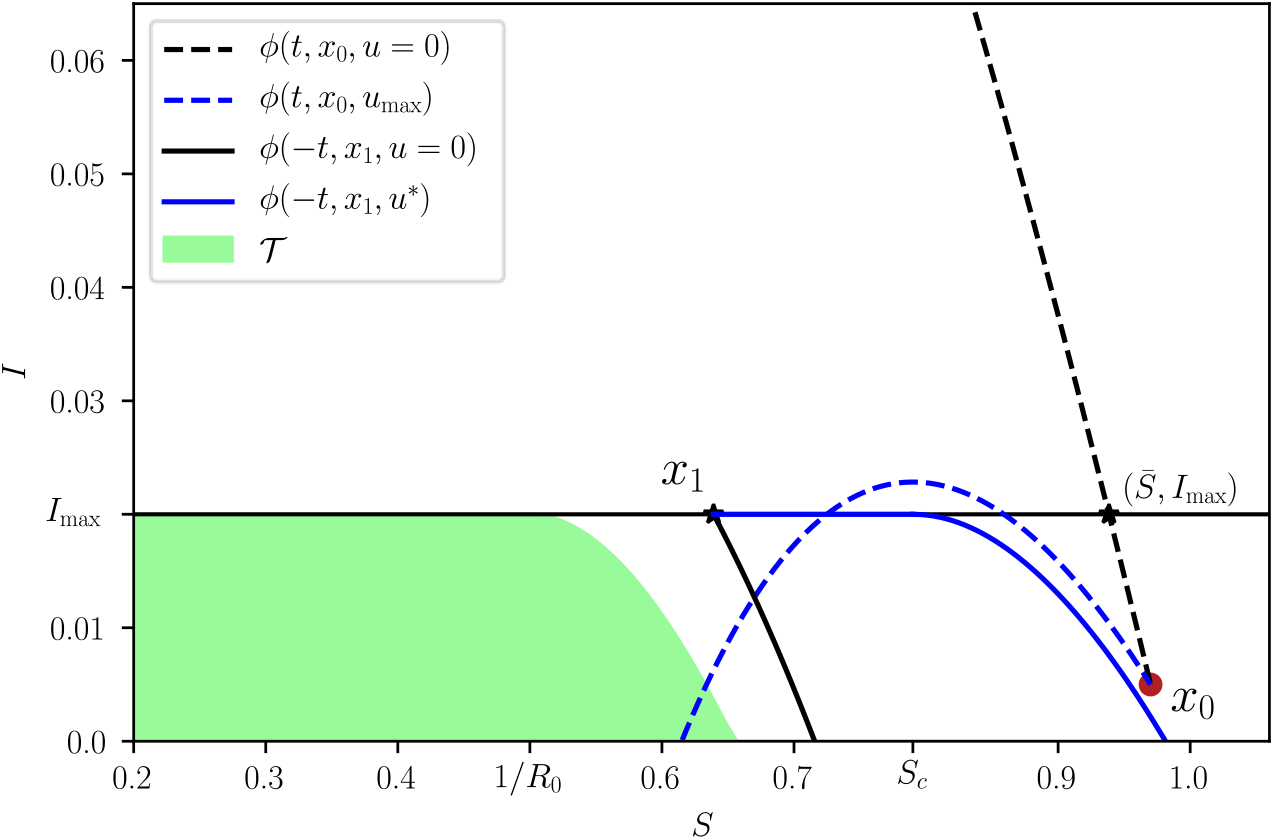
For the initial condition *x*_0_ the problem is unfeasible, *I*_max_ will be surpassed, no matter which strategy is used. The figure was generated with *R*_0_ = 2, *R*_*c*_ = 1.27 and *I*_max_ = 0.02.

**Supplementary Figure S8.**
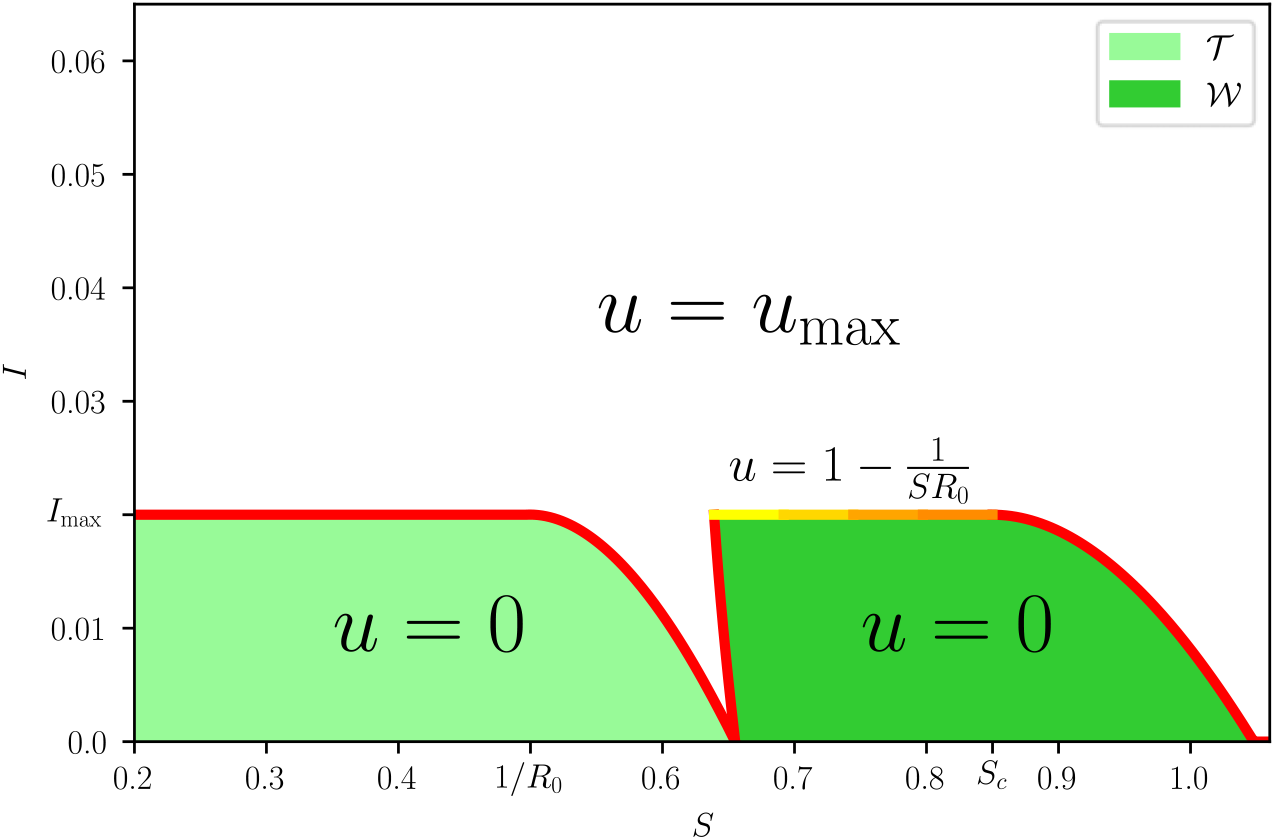
The optimal feedback strategy (S5). The target and waiting sets, 𝒯 and 𝒲, are illustrated for *R*_0_ = 2, *R*_*c*_ = 1.18 and *I*_max_ = 0.02.

The optimal control feedback is thus given by

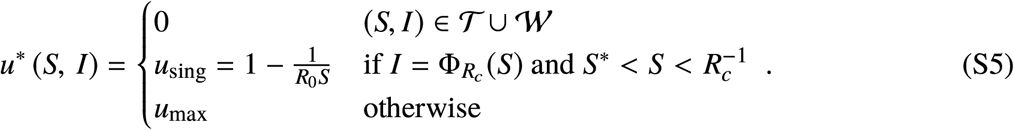

Such strategy is summarized in Fig. S8.

Alternatively, we can implement a pure switching control since the “equivalent control”[2] will realize the singular control on the singular arc,

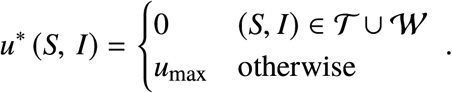

Note that this control strategy extends the control action beyond the region where the optimal control is feasible.

### S2. Necessary and sufficient conditions for the existence of optimal NPIs

Let (*S*_0_, *I*_0_) denote the initial state of the SI model. As shown in Supplementary Note S1, the necessary and sufficient condition for the existence of NPIs is that 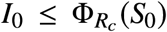 where 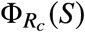 is the separating curve. To characterize a condition that is independent of the initial state, we consider the limit case of *S*_0_ = 1 and *I*_0_ = 0. Under this assumption, the necessary and sufficient condition of existence is that 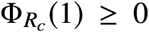. In other words, the boundary of existence of NPIs is when the separating curve exactly crosses *I* = 0 at *S* = 1. Substituting *S* = 1 in the separating curve we obtain the condition

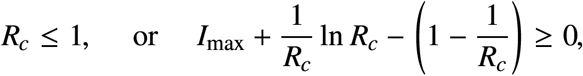

which is precisely the inequality (1) of the Main Text..

### S3. Robustness of the optimal intervention

Here we describe the models used to evaluate the robustness of the optimal intervention.

#### S3.1 Robustness to the presence of demography and an incubation period

To evaluate the robustness of the optimal intervention to the presence of an incubation period of the disease, we considered the SEIR dynamics

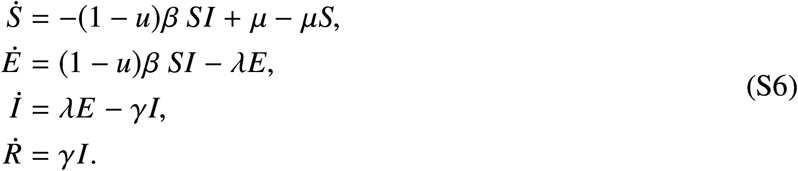

Above, *E*(*t*) denotes the fraction of individuals in the population exposed to the disease, but which are not yet infectious, at time *t*. The parameter 1/*λ* ≥ 0 denotes the *incubation period* of the disease in units of days. The parameter *μ* ≥ 0 denotes the *recruitment rate* in units of days^−1^. For the result of our paper, we choose *μ* = 1/(365 · 75) corresponding to a life expectancy of 75 years. For this model, the intervention we apply is *u*(*t*) = *u*^∗^(*S*(*t*), *I*(*t*)) with *u*^∗^(*S, I*) as in Eq. (S5).

#### S3.2 Robustness to the presence of hidden infected individuals

To evaluate the robustness of the optimal intervention to hidden infected individuals, consider that that infections can be symptomatic or asymptomatic. We assume that all asymptomatic infections do not require hospital care, and hence remain undetected by the epidemic surveillance system. To model this scenario, we consider the dynamics

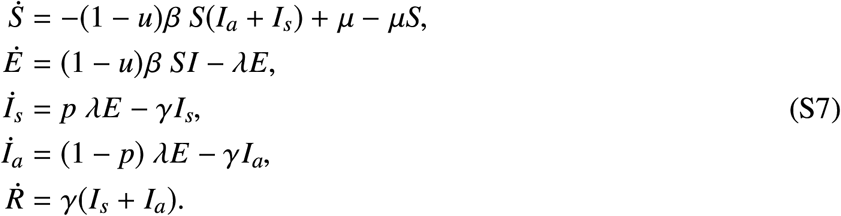

Above, *I*_*s*_ denotes the fraction of symptomatic infections and *I*_*a*_ the fraction of asymptomatic ones. The model assumes that a fraction *p* ∈ [0, 1] of exposed individuals result in symptomatic infections, and the rest (1− *p*) in asymptomatic ones. We assume that infectious period 1/*γ* is the same for both symptomatic and asymptomatic individuals. For the results of our paper, we choose *λ* = 1/7. Since we assume that only symptomatic individuals end up requiring hospital care, we consider that the objective is to keep *I*_*s*_(*t*) ≤ *I*_max_ only. The control applied is *u*(*t*) = *u*^∗^(*S*(*t*), *I*_*s*_(*t*)) where *u*^∗^(*S, I*) is given by Eq. (S5).

### S4. Application to the COVID-19 pandemic

#### S4.1 Estimate for the fraction of infected individuals requiring intensive care

For COVID-19 pandemic by the SARS-CoV-2 virus, we estimated the fraction *f* of infected individuals requiring intensive-care under the following assumptions:

1. Current estimates for the fraction *p* ∈ [0, 1] of infected individuals that are symptomatic show a large variability [3], ranging from a 20/100 in a report of the World Health Organization, to 96/100 in a study of 328 adults in Shanghai[4]. We take the nominal value of *p*_0_ = 60/100.
2. Following Kremer et al.[5], we assume that from the individuals that are symptomatic, a fraction 15/100 develop severe symptoms.
3. Finally, following Li et al. [6], from the individuals that develops severe symptoms, we assume that the fraction 28/100 will require intensive care.

Under the above assumptions, the fraction of infected individuals requiring intensive care has a nominal value

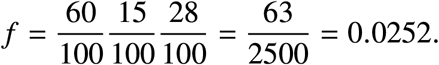

#### S4.2 Data used in our analysis

Supplementary Fig. S9 shows the data used for our analysis. Data was collected using the following methodology:

1. **Number of intensive care beds in each city**. This was obtained from official statements when possible (e.g., the Massachusetts Department of Public Health for Boston). In other cases, this number was obtained from public statements of authorities of each city. A complete list of the references appears in the Supplementary Fig. S9.
2. **Population in each city**. Data was obtained from Wikipedia.
3. **Reduction of mobility in each city**. This was obtained from Google Community Mobility Reports https://www.google.com/covid19/mobility/. For our analysis, we considered three categories of mobility: retail & recreation, transit stations, and workplaces. To estimate an overall mobility reduction, we averaged the mobility reduction in these three categories from March 19 to April 30. Data was accessed on May 7, 2020.
4. **Basic reproduction number**. We estimated this quantity from the value of the effective time-varying reproduction number *R*_*t*_ at the start of the pandemic around March 8, 2020. These estimates were obtained from the website https://epiforecasts.io/covid/.

**Supplementary Figure S9.**
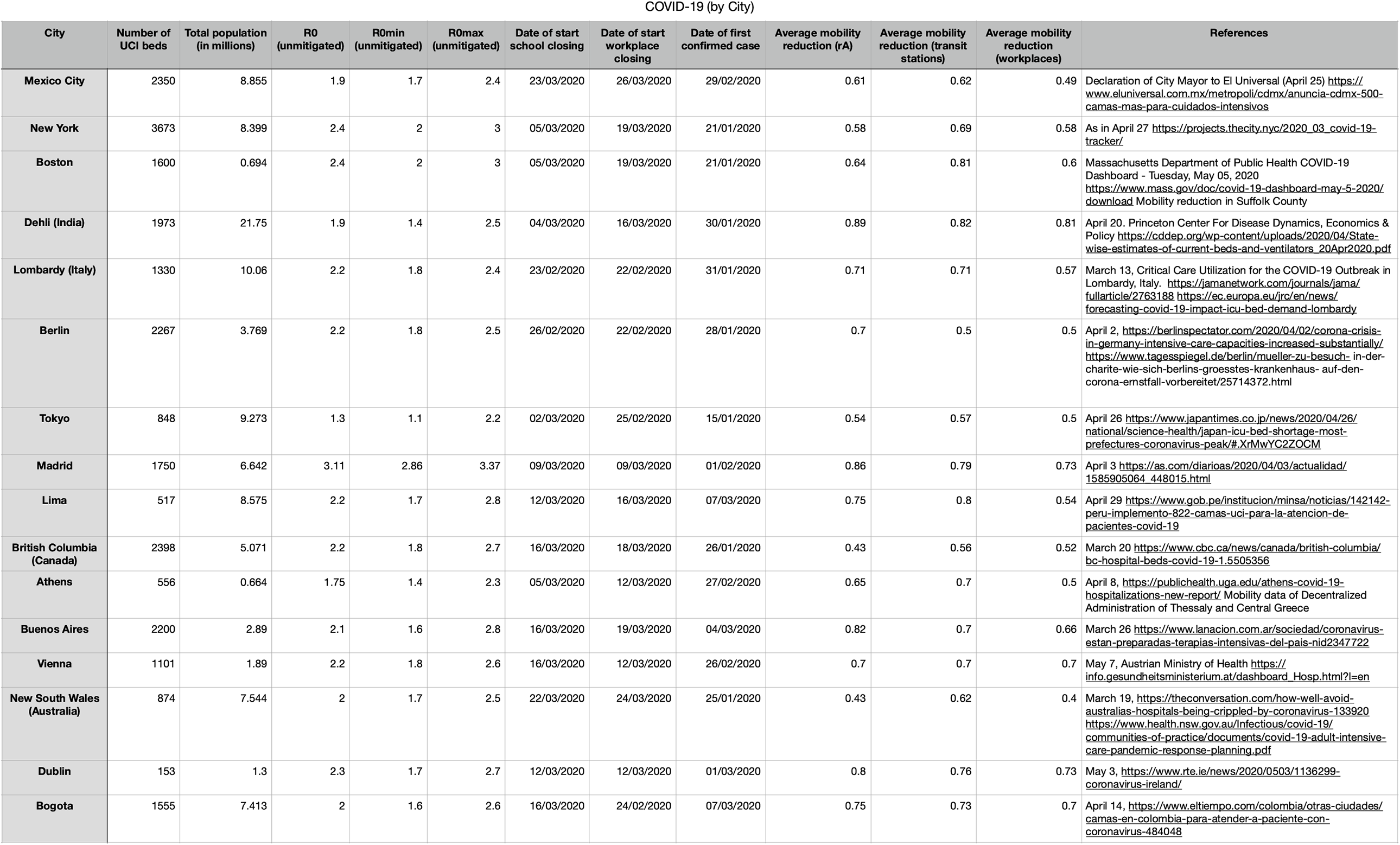
Table with the response of 16 cities during the COVID-19 pandemic.

### S5. Related work

For the control of infectious diseases, there is a large body of work using optimal control methods to design interventions, including vaccination and quarantines[7, 8], drug treatments[9], or dispersal of insecticides and education campaigns[10]. The standard tool to solve these optimal control problem is the celebrated Pontryagin’s Maximum Principle[11]. However, note that the Maximum Principle only gives necessary conditions for optimality. The gap between the necessary and sufficient conditions for optimality needs to be closed using additional arguments, often relying on assuming that the control appears multiplying an affine function of the state variables. This assumption is not satisfied in our formulation of optimal NPIs. We emphasize that the optimal interventions obtained from this approach result in *open loop* strategies which only depend on time. By contrast, our analysis gives a feedback optimal strategy that characterizes the optimal action to make according to the actual state of the epidemic. Indeed, our characterization of optimal NPIs does not rely on the Maximum Principle. Instead, the low dimensional of our model allows us to apply Green’s Theorem to compare the cost of two Different interventions. The consequence of our approach is that we obtain a feedback or *closed loop* strategy that corrects itself based on the actual state of the epidemic.

The COVID-19 pandemic has stirred much interest on designing non-pharmaceutical interventions. This has led to strategies like interspacing mitigation with brief periods of activity[12]. Optimal control methods have been also applied, for example to minimize the peak of infection[13], minimize the number of infections[14], minimize the economic costs[15], or maximize welfare[16]. Compared to these studies, our analytical characterization of optimal NPIs provides gives us a complete understanding of the optimal decisions that need to be made. For example, no intervention is needed before reaching the separating curve.

## Footnotes

1 If we select *S* > *S*_0_ the obtained value of *I* is reached in a past time (*t* < 0).

## Notes

### Competing Interest Statement

The authors have declared no competing interest.

### Funding Statement

M.T.A. gratefully acknowledges the financial support from CONACyT project A1-S-13909, México. J. X.V.H. acknowledges the financial support from UNAM IN115720.

### Summary of Updates

Revised calculations

